# Metal mixtures associate with higher amyotrophic lateral sclerosis risk and mortality independent of genetic risk and correlate to self-reported exposures: a case-control study

**DOI:** 10.1101/2024.02.27.24303143

**Authors:** Dae Gyu Jang, John Dou, Emily J. Koubek, Samuel Teener, Lili Zhao, Kelly M. Bakulski, Bhramar Mukherjee, Stuart A. Batterman, Eva L. Feldman, Stephen A. Goutman

**Affiliations:** Department of Neurology, University of Michigan, Ann Arbor, MI; NeuroNetwork for Emerging Therapies, University of Michigan, Ann Arbor, MI; Department of Epidemiology, University of Michigan, Ann Arbor, MI; Department of Biostatistics, Corewell Health, Royal Oak, MI; Department of Biostatistics, University of Michigan, Ann Arbor, MI; Department of Environmental Health Sciences, University of Michigan, Ann Arbor, MI

**Keywords:** Amyotrophic lateral sclerosis (ALS), metals, polygenic risk score, environmental risk score

## Abstract

**Background:** The pathogenesis of amyotrophic lateral sclerosis (ALS) involves both genetic and environmental factors. This study investigates associations between metal measures in plasma and urine, ALS risk and survival, and exposure sources.

**Methods:** Participants with and without ALS from Michigan provided plasma and urine samples for metal measurement via inductively coupled plasma mass spectrometry. Odds and hazard ratios for each metal were computed using risk and survival models. Environmental risk scores (ERS) were created to evaluate the association between exposure mixtures and ALS risk and survival and exposure source. ALS (ALS-PGS) and metal (metal-PGS) polygenic risk scores were constructed from an independent genome-wide association study and relevant literature-selected SNPs.

**Results:** Plasma and urine samples from 454 ALS and 294 control participants were analyzed. Elevated levels of individual metals, including copper, selenium, and zinc, significantly associated with ALS risk and survival. ERS representing metal mixtures strongly associated with ALS risk (plasma, OR=2.95, CI=2.38-3.62, *p*<0.001; urine, OR=3.10, CI=2.43-3.97, *p*<0.001) and poorer ALS survival (plasma, HR=1.42, CI=1.24-1.63, *p*<0.001; urine, HR=1.52, CI=1.31-1.76, *p*<0.001). Addition of the ALS-PGS or metal-PGS did not alter the significance of metals with ALS risk and survival. Occupations with high potential of metal exposure associated with elevated ERS. Additionally, occupational and non-occupational metal exposures associated with measured plasma and urine metals.

**Conclusion:** Metals in plasma and urine associated with increased ALS risk and reduced survival, independent of genetic risk, and correlated with occupational and non-occupational metal exposures. These data underscore the significance of metal exposure in ALS risk and progression.

## INTRODUCTION

The neurodegenerative condition amyotrophic lateral sclerosis (ALS) results in motor impairment affecting cranial, trunk, limb, and respiratory skeletal muscles.^1^ Currently, no cure exists for ALS, and symptoms progressively worsen, leading to a survival of 2-4 years. ALS pathogenesis is multifaceted and the gene-time-environment hypothesis proposes that ALS is triggered by environmental exposures acting upon a background of genetic susceptibility.^2^ In our recent study involving ALS patients in Michigan, we found that polygenic risk contributes to the development of ALS.^3^ Additionally, we demonstrated that environmental risk scores (ERS), representing various persistent organic pollutants (POPs) simultaneously, associate with ALS risk and survival.^4–6^ Thus, both genetic and environmental factors contribute to ALS risk and progression in our Michigan ALS cohort.

Self-reported occupational exposures to metals are linked to increased ALS risk,^7^ and metal exposure is widely acknowledged as a contributing factor to the disease.^8^ Therefore, we investigated the impact of metal levels in plasma and urine, individually and as a mixture modeled by ERS, on ALS risk and survival. By analyzing metal levels in both blood and urine samples, we aimed to encompass the diverse sources and exposure pathways of metals, as well as the biotransformation within and elimination processes from the body,^9^ thus capturing a more comprehensive picture of metal exposure. Furthermore, we developed polygenic risk scores (PGS) representing genes associated with both ALS risk and metal metabolism to explore the potential interplay between genetic background and metal exposure in ALS development. Finally, we examined the correlations between metal mixtures and self-reported occupational exposures. These findings provide valuable insight into underlying disease mechanisms and highlight potentially modifiable risk factors for ALS.

## METHODS

### Participants and biosamples

Details of this cohort are previously published.^3–7, 10–14^ Briefly, ALS participants were enrolled at the University of Michigan Pranger ALS Clinic. All ALS patients meeting Gold Coast criteria able to communicate in English were approached for participation. Control participants were identified via population outreach using a University of Michigan research database, random address mailing, and Facebook advertisements. All participants provided informed consent and the study was IRB approved at the University of Michigan (HUM28826). Participants were required to provide biosamples for metals analyses to be eligible for inclusion in this study.

Plasma samples were collected between February 26, 2012 - December 7, 2022 and urine samples between September 23, 2015 - September 9, 2022. Information abstracted from medical records included use of supplements and/or vitamins and disease characteristics (e.g., onset segment, age of diagnosis) for ALS cases. *C9orf72* gene status was obtained from medical records and previously published data.^3^

Peripheral blood was collected via peripheral venipuncture, processed for plasma, aliquoted into cryovials, and stored at -80°C. Urine was collected in a specimen collection cup, aliquoted into cryovials, and stored at -80°C. Samples were shipped on dry ice to the Dartmouth Trace Element Analysis Core for analysis following their standard protocols. Briefly, plasma and urine samples were homogenized and then acid-digested with 1% HNO_3_. The following 23 metals were measured in plasma and urine samples: aluminum, antimony, arsenic, barium, beryllium, cadmium, chromium, cobalt, copper, iron, lead, manganese, mercury, molybdenum, nickel, selenium, silver, strontium, thallium, tin, uranium, vanadium, and zinc. Analysis was performed by inductively coupled plasma mass spectrometry (ICP-MS) on an Agilent 8900 ICP-MS. Five or more multi-point calibration curves were constructed for each element with a correlation coefficient criteria>0.995. Following calibration, an initial calibration blank and an initial calibration verification were performed. Continuing calibration verifications were completed every 10 samples.

### Polygenic scores

Two polygenic risk scores (PGS) were used in this study cohort. The first, an ALS-PGS was constructed as previously reported^3^ but updated with weights from a recent large-scale ALS genome-wide association study (GWAS, **Supplemental Table 1**).^15^ Briefly, PRSice-2 software was employed to clump and prune SNPs using default settings, followed by the application of weights from the parent GWAS to construct the ALS-PGS. Notably, the ALS-PGS was exclusively constructed for participants not included in the large ALS GWAS. Additionally, a metal-PGS was constructed to represent metabolism of metals (metal-PGS), utilizing a selection of relevant SNPs identified from literature (**Supplemental Table 2**). As the selected SNPs originated from various studies with diverse outcomes, weights were not applied based on effect estimates; instead, the metal-PGS was constructed based on the count of risk alleles.

### Genetic samples

Samples were genotyped in three separate batches. In the first batch, 188 samples were genotyped on the OmniExpress array. In a second batch, as previously described,^3^ 512 samples were genotyped using the Infinium Multi-Ethnic Global-8 array. Lastly, 305 samples were genotyped using the Infinium Global Diversity Array-8 Kit. For each batch of data, QC filtering was applied at the SNP (**Supplemental Figure 1**) and sample level (**Supplemental Figure 2**). A total of 926 samples were available after QC and merging. These were then imputed to the Haplotype Reference Consortium reference panel via the University of Michigan imputation server. After filtering the SNPs that had imputation Rsq<0.5 and minor allele frequency<1%, 7,733,379 SNPs remained (**Supplemental Figure 1**).

### Missing data and multiple imputation

Measurements of metals in plasma and urine are subject to missingness due to detection limits and other issues. Five metals that were found below the limit of detection (LOD) in over 40% of samples were excluded from ERS construction. Certain metals were also manually excluded from ERS construction due to the unsuitability of the biofluid source for accurate and representative measurement.^9^ Metals excluded from the plasma ERS due to biofluid unsuitability included arsenic,^16^ cadmium,^9^ and silver.^17^ Metals excluded from urine ERS included iron, manganese,^18^ silver,^17^ and strontium.^19^ Metal concentrations not excluded but otherwise missing due to non-detection were imputed by *LOD* √2.

Statistical models were adjusted for covariates. Covariates subject to missingness included education, military service, family history of ALS, age at diagnosis, onset segment, and El Escorial criteria. To handle missing data, multiple imputation by chained equations was used and stratified by case-control status for each sample source. The imputation model for controls included metals (log-transformed), age at sample collection, sex, military service, and family history of ALS. The imputation model for cases included all predictors in the control model, as well as age at diagnosis, onset segment, El Escorial criteria, initial ALSFRS-R score, smoking status, education, and cumulative hazard rate (Nelson-Aalen estimator) from diagnosis to last follow-up. Twenty multiple imputation datasets were generated using a predictive mean matching model.

### Descriptive statistics

Descriptive statistics summarized metal level distributions. Spearman correlation coefficients were employed to visualize correlation plots. Statistics, including means, standard deviations (SDs), 10th, 25th, 50th, 75th, and 90th quantiles, minimum and maximum of the distributions of metal levels in ALS cases and controls were computed. Wilcoxon rank sum tests were conducted to compare metal levels in ALS cases and controls. *P*-values were adjusted for multiple comparisons by Benjamini-Hochberg correction.

### Risk model

Logistic regression models were constructed to examine the relationships between individual metals and ALS risk. Adjustment covariates were age at sample collection, sex, and military service. Models were fitted independently across 20 imputation datasets and results across models were pooled using Rubin’s rule. A Benjamini-Hochberg correction was applied to address the issue of multiple of comparisons.

To assess multiple metals concurrently, environmental risk scores (ERS) were constructed. As metal levels differ between plasma and urine samples, separate ERS scores for plasma (ERS ^PR^; plasma risk ERS) and urine (ERS^UR^; urine risk ERS) were developed. An adaptive elastic net approach applied to the combined 20 imputed datasets using the R-package *miselect*^20^ determined the ERS weights. To optimize the performance of the adaptive elastic net, hyperparameters were carefully tuned through a 5-fold cross-validation process with the aim of minimizing deviance, based on the log likelihood for a logistic regression model.

A logistic regression model was used to evaluate the interaction between plasma/urine levels and ALS-/metal-PGS. In this model, adjusted covariates included age at sample collection, sex, military service, and the first five genetic principal components.^3^ Of note, *C9orf72* gene positivity status was not adjusted for in the risk model as there were no *C9orf72* positive cases in the control population.^3^

### Survival model

Associations between survival time since ALS diagnosis and individual metal exposures were estimated using a Cox proportional hazards model. Covariates included age at diagnosis, sex, family history of ALS, bulbar vs non-bulbar onset, diagnostic El Escorial criteria, and time between symptom onset and diagnosis. Inferences regarding each metal were adjusted for multiple comparisons using a Benjamini-Hochberg correction.

Survival ERS was constructed via application of a Cox regression with ridge penalty to the combined twenty imputation datasets, following published methods.^4, 5^ Again, separate ERS were developed for plasma (ERS^PS^; plasma survival ERS) and urine (ERS^US^; urine survival ERS). The resulting ERS represents the association between metal profile and survival time. Hyperparameters of the ridge penalty were tuned through a 5-fold cross-validation process with the aim of minimizing the deviance based on the log partial likelihood for a Cox model. Kaplan-Meier survival curves were utilized to depict the survival functions of quartiles based on ERS. Survival curves were adjusted for covariates via the inverse probability weights method.Propensity scores were computed using the multinomial logistic regression models, with ERS quartiles as outcome variables and adjustment covariates (as stated above) as predictors.

Interactions between genetic (ALS/metal PGS) and environmental factors (plasma/urine metal levels) were also assessed utilizing a Cox survival model. The model was adjusted for age at diagnosis, sex, family history of ALS, bulbar vs. non-bulbar onset, initial El-Escorial criteria, time between symptom onset and diagnosis, the first five genetic principal components, and *C9orf72* gene positivity.

### Occupations and self-reported occupational and non-occupational exposures

Occupational^7^ and non-occupational^21^ data were generated as previously described. Briefly, participants provided a list of prior job titles and responsibilities that were used to generate standard occupational classification (SOC) codes (**Supplemental Table 3**). Participants also answered questions regarding known exposures to metals in their most recent occupation and in non-occupational settings. SOC codes were separated into high- and low-metal exposure groups based on previously published results.^7^ Distributions of ERS^PR^ and ERS^UR^ by high/low metal exposure occupational groups were compared using Wilcoxon tests. Associations between occupational/non-occupational metal exposures and urine/plasma metals were assessed using the partial least squares method feature in the R-package “mixOmics”^22^ and estimated associations were visualized in heatmap plots.

### Statistical software

All analyses were performed using R statistical software, version 4.2.3 (https://www.R-project.org/).

## RESULTS

### Participants and metals analysis

The full population cohort included 454 ALS patients and 295 control participants, with plasma samples analyzed for 387 ALS patients and 281 control participants (**Table 1**, **Figure 1**), and urine samples obtained from 322 ALS patients and 194 control participants (**Table 2**, **Figure 1**). In the plasma cohort, ALS individuals were older (median age 65 vs. 62 years, *p*<0.001) and had a higher proportion of males (56% vs. 48%, *p*=0.036) than control participants (**Table 1**).

**Figure 1.**
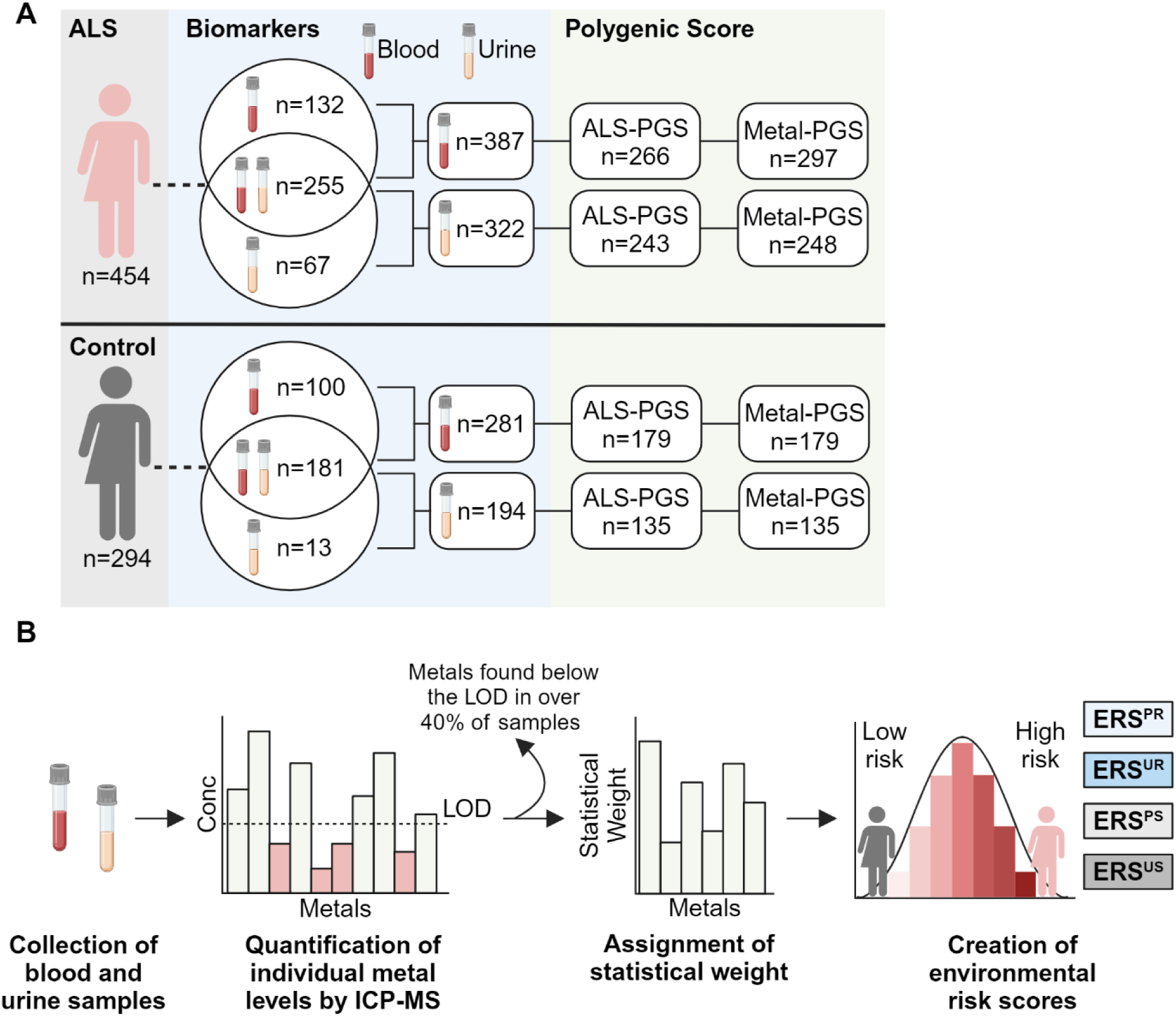
Study overview. **(A)** Participants with (n=454) or without (n=294) amyotrophic lateral sclerosis (ALS) provided blood and/or urine samples. Polygenic risk scores (PGS) for ALS risk or genes related to metal metabolism were available for a subset of participants. **(B)** Blood and urine samples were collected from study participants and analyzed by inductively coupled plasma mass spectrometry (ICP-MS) to measure the concentrations of various metals. After removing metals in which over 40% of the samples fell under the limit of detection (LOD), the statistical weight of each metal was determined, and overall environmental risk scores (ERS) were calculated. Individual ERS for plasma and urine samples were developed for risk (plasma, ERS^PR^; urine, ERS^UR^) and survival (plasma, ERS^PS^; urine, ERS^US^). These scores represent the cumulative exposure to various metals and serve as a comprehensive indicator of environmental risk. Image created using BioRender.com.

**Table 1.** ALS and control participant demographics: Plasma.

| Covariate | ALS, N = 387<sup>1</sup> | Control, N = 281<sup>1</sup> | p-value<sup>2</sup> |
| --- | --- | --- | --- |
| Age at Sample Collection | 65 (58-73) | 62 (55-68) | **<0.001** |
| Sex |  |  | **0.038** |
| Female | 171 (44%) | 147 (52%) |  |
| Male | 216 (56%) | 134 (48%) |  |
| Ethnicity |  |  | **0.036** |
| Hispanic or Latino | 4 (1.0%) | 9 (3.4%) |  |
| Not Hispanic or Latino | 378 (99%) | 255 (97%) |  |
| Missing | 5 | 17 |  |
| Race |  |  | 0.2 |
| American Indian and Alaska native | 2 (0.5%) | 0 (0%) |  |
| Asian | 1 (0.3%) | 4 (1.4%) |  |
| Black or African American | 13 (3.4%) | 10 (3.6%) |  |
| White or Caucasian | 370 (96%) | 267 (95%) |  |
| Missing | 1 | 0 |  |
| Military Service |  |  | 0.2 |
| Yes | 46 (12%) | 22 (8.9%) |  |
| No | 326 (88%) | 224 (91%) |  |
| Missing | 15 | 35 |  |
| Education |  |  | **<0.001** |
| High school or less | 109 (29%) | 13 (5.3%) |  |
| Bachelor's degree | 75 (20%) | 93 (38%) |  |
| Some postsecondary | 125 (34%) | 51 (21%) |  |
| Graduate degree | 63 (17%) | 90 (36%) |  |
| Missing | 15 | 34 |  |
| Family History of ALS | 42 (11%) | 0 (0%) | **<0.001** |
| Missing | 12 |  |  |
| Age at Diagnosis | 65 (57-72) |  |  |
| Missing | 2 |  |  |
| Diagnosis |  |  |  |
| ALS | 358 (93%) |  |  |
| ALS/Frontotemporal dementia | 17 (4.4%) |  |  |
| Brachial amyotrophic diplegia | 6 (1.6%) |  |  |
| Flail leg | 2 (0.5%) |  |  |
| Progressive muscular atrophy | 4 (1.0%) |  |  |
| EI Escorial Criteria |  |  |  |
| Definite | 114 (31%) |  |  |
| Probable | 114 (31%) |  |  |
| Probable, lab supported | 100 (27%) |  |  |
| Possible | 42 (11%) |  |  |
| Suspected | 0 (0%) |  |  |
| Missing | 17 |  |  |
| Onset Segment |  |  |  |
| Bulbar | 99 (26%) |  |  |
| Cervical | 126 (33%) |  |  |
| Lumbar | 152 (39%) |  |  |
| Respiratory | 4 (1.0%) |  |  |
| Thoracic | 4 (1.0%) |  |  |
| Missing | 2 |  |  |
| ALSFRS-R | 37 (33-41) |  |  |
| Missing | 2 |  |  |
| Time Between Symptom Onset and Diagnosis (Years) | 1.04 (0.64-1.77) |  |  |
| Missing | 5 |  |  |
| Time Between Diagnosis and Sample (Years) | 0.58 (0.27-1.13) |  |  |
| Missing | 2 |  |  |

<sup>1</sup>Median (25%-75%); n (%)
<sup>2</sup>Wilcoxon rank sum test; Pearson's Chi-squared test; Fisher's exact test
Abbreviations: ALS, amyotrophic lateral sclerosis; ALSFRS-R, Revised ALS Functional Rating Scale

**Table 2.**
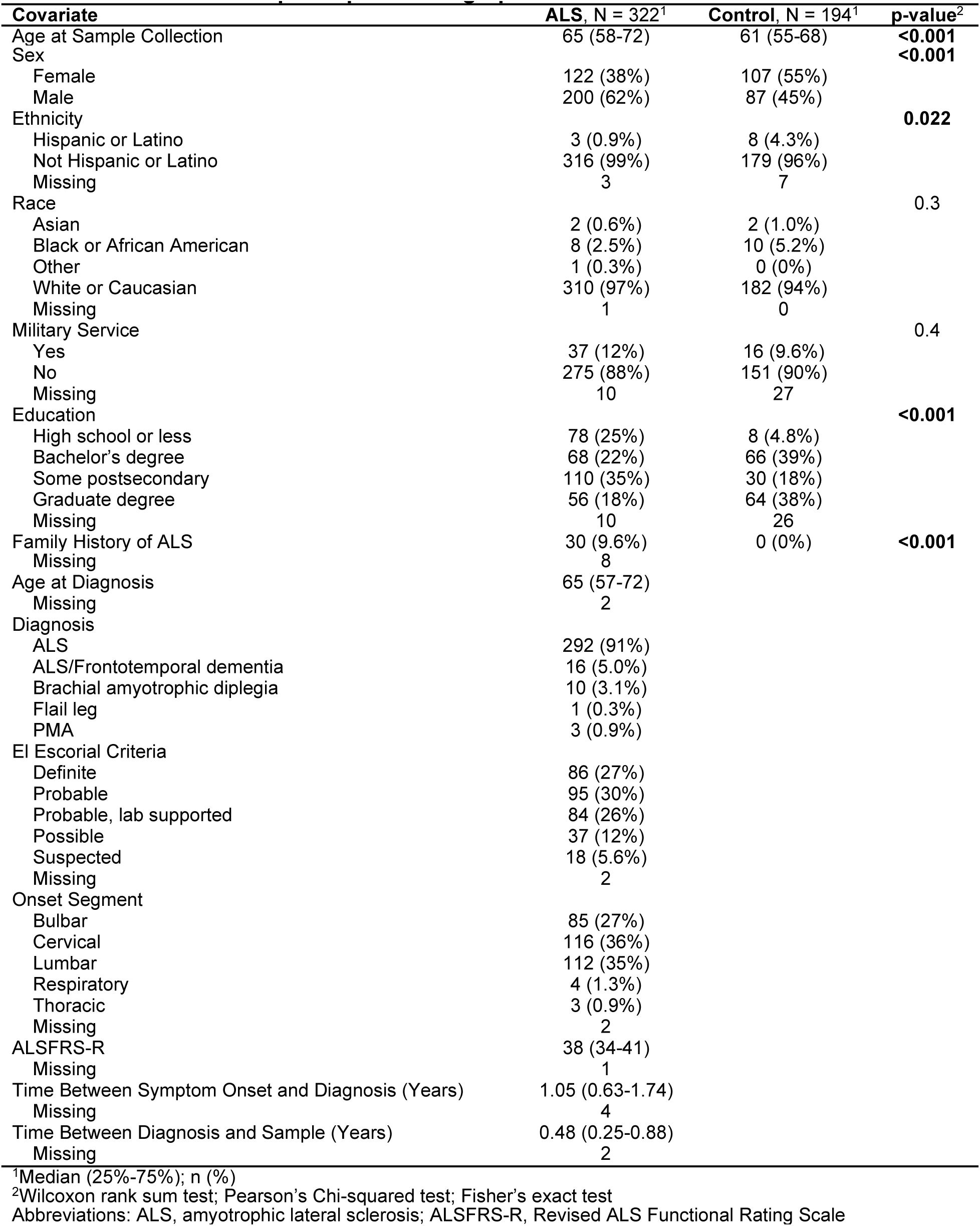
ALS and control participant demographics: Urine.

| **Covariate** | **ALS, N = 322<sup>1</sup>** | **Control, N = 194<sup>1</sup>** | **p-value<sup>2</sup>** |
| --- | --- | --- | --- |
| Age at Sample Collection | 65 (58-72) | 61 (55-68) | **<0.001** |
| Sex |  |  | **<0.001** |
| Female | 122 (38%) | 107 (55%) |  |
| Male | 200 (62%) | 87 (45%) |  |
| Ethnicity |  |  | **0.022** |
| Hispanic or Latino | 3 (0.9%) | 8 (4.3%) |  |
| Not Hispanic or Latino | 316 (99%) | 179 (96%) |  |
| Missing | 3 | 7 |  |
| Race |  |  | 0.3 |
| Asian | 2 (0.6%) | 2 (1.0%) |  |
| Black or African American | 8 (2.5%) | 10 (5.2%) |  |
| Other | 1 (0.3%) | 0 (0%) |  |
| White or Caucasian | 310 (97%) | 182 (94%) |  |
| Missing | 1 | 0 |  |
| Military Service |  |  | 0.4 |
| Yes | 37 (12%) | 16 (9.6%) |  |
| No | 275 (88%) | 151 (90%) |  |
| Missing | 10 | 27 |  |
| Education |  |  | **<0.001** |
| High school or less | 78 (25%) | 8 (4.8%) |  |
| Bachelor's degree | 68 (22%) | 66 (39%) |  |
| Some postsecondary | 110 (35%) | 30 (18%) |  |
| Graduate degree | 56 (18%) | 64 (38%) |  |
| Missing | 10 | 26 |  |
| Family History of ALS | 30 (9.6%) | 0 (0%) | **<0.001** |
| Missing | 8 |  |  |
| Age at Diagnosis | 65 (57-72) |  |  |
| Missing | 2 |  |  |
| Diagnosis |  |  |  |
| ALS | 292 (91%) |  |  |
| ALS/Frontotemporal dementia | 16 (5.0%) |  |  |
| Brachial amyotrophic diplegia | 10 (3.1%) |  |  |
| Flail leg | 1 (0.3%) |  |  |
| PMA | 3 (0.9%) |  |  |
| El Escorial Criteria |  |  |  |
| Definite | 86 (27%) |  |  |
| Probable | 95 (30%) |  |  |
| Probable, lab supported | 84 (26%) |  |  |
| Possible | 37 (12%) |  |  |
| Suspected | 18 (5.6%) |  |  |
| Missing | 2 |  |  |
| Onset Segment |  |  |  |
| Bulbar | 85 (27%) |  |  |
| Cervical | 116 (36%) |  |  |
| Lumbar | 112 (35%) |  |  |
| Respiratory | 4 (1.3%) |  |  |
| Thoracic | 3 (0.9%) |  |  |
| Missing | 2 |  |  |
| ALSFRS-R | 38 (34-41) |  |  |
| Missing | 1 |  |  |
| Time Between Symptom Onset and Diagnosis (Years) | 1.05 (0.63-1.74) |  |  |
| Missing | 4 |  |  |
| Time Between Diagnosis and Sample (Years) | 0.48 (0.25-0.88) |  |  |
| Missing | 2 |  |  |

<sup>1</sup>Median (25%-75%); n (%)<sup>2</sup>Wilcoxon rank sum test; Pearson's Chi-squared test; Fisher's exact test
Abbreviations: ALS, amyotrophic lateral sclerosis; ALSFRS-R, Revised ALS Functional Rating Scale

Similarly, the urine cohort showed comparable trends, with ALS patients being older (median age 65 vs. 61 years, *p*<0.001) and more frequently male (62% vs. 45%, *p*<0.001) than controls (**Table 2**). Educational attainment was higher among controls in both cohorts. Furthermore, ALS characteristics were consistent between plasma and urine cohorts and were typical, with 93% and 91% of participants presenting with classical ALS, 31% and 27% meeting El Escorial Definite ALS criteria, and 26% and 27% having bulbar onset, respectively (**Tables 1 and 2**). Samples of both plasma and urine were available for 255 ALS patients and 181 controls (**Supplemental Table 4**).

Concentrations of the 23 metals detected in plasma and urine samples are listed in **Supplemental Tables 5 and 6**, respectively. Metals found below the LOD in over 40% of plasma samples included aluminum, beryllium, mercury, molybdenum, and uranium, while in urine samples, these were beryllium, chromium, manganese, mercury, and silver. Among plasma metals, levels of cadmium, copper, lead, selenium, vanadium, and zinc were significantly (*p_adjusted_*<0.05) higher in ALS patients than control participants, while antimony, arsenic, barium, and strontium levels were higher in control individuals (**Supplemental Table 5**). In urine, concentrations of aluminum, barium, cadmium, copper, iron, molybdenum, selenium, strontium, tin, uranium, vanadium, and zinc were significantly higher in ALS patients than control participants (**Supplemental Table 6**). **Supplemental Figure 3** shows a heat map of Spearman correlations between metal biomarkers, indicating more pronounced correlations among urine metals compared to plasma metals, with few metals showing significant interclass correlation between plasma and urine measures.

### ALS risk

#### Plasma

Odds ratios (ORs) for metals exhibited a wide range, spanning from 0.58 for antimony to 1.58 for selenium (**Figure 2**). Analyzing the entire population with a significance threshold of *p_adjusted_*<0.05, several plasma metals demonstrated significant associations with an elevated risk of ALS, including copper, lead, selenium, and zinc. Conversely, antimony, barium, nickel, and strontium were linked with a reduced risk of ALS. Given the potential exposure to multiple metals over a lifetime, an ERS for metal mixtures was constructed to assess the relationship between metal combinations and ALS risk. Metals included in the ERS^PR^ are detailed in **Table 3**. Analysis revealed a significant association between the ERS^PR^ and ALS risk (**Supplemental Figure 4A**), with a one standard deviation (SD) increase in ERS^PR^ based on the control population corresponding to an OR=2.95 (*p*<0.001, **Figure 2**).

**Figure 2.**
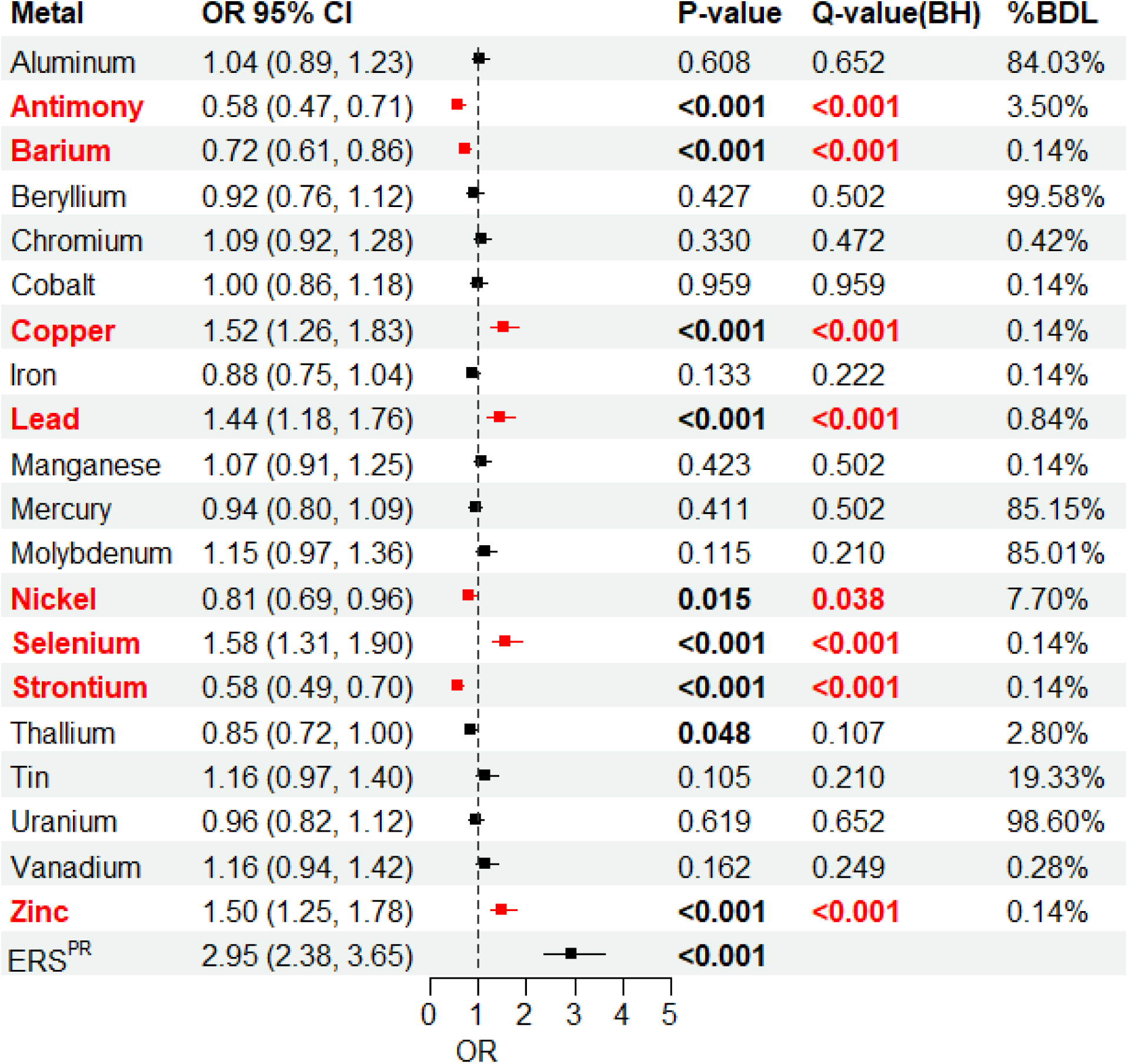
Plasma single metal and mixture associations with ALS risk. Single metal logistic regression models based on plasma samples where the outcome is case/control status, the predictors are log-transformed, standardized metal levels, and the covariates are continuous age at sample collection, sex, and military service. Mixture metal ERS^PR^ (plasma risk environmental risk score). %BDL, percentage of samples below detection limit; ALS, amyotrophic lateral sclerosis; BH, Benjamini-Hochberg, red font are significant correlations; CI, confidence interval; OR, odds ratio corresponding to one standard deviation increase in log-transformed metals.

**Table 3.** Metals included in environmental risk scores (ERS)

| Metal | Weights |  |  |  |
| --- | --- | --- | --- | --- |
|  | ERS<sup>PR</sup> | ERS<sup>PS</sup> | ERS<sup>UR</sup> | ERS<sup>US</sup> |
| Aluminum |  |  | 0.000 | 0.014 |
| Antimony | -0.352 | 0.022 | 0.000 | 0.003 |
| Arsenic |  | -0.022 | -0.103 | -0.007 |
| Barium | -0.251 | 0.000 | 0.188 | 0.003 |
| Cadmium |  | 0.022 | 0.144 | 0.021 |
| Chromium | 0.192 | -0.015 |  |  |
| Cobalt | 0.056 | -0.006 | -0.023 | 0.003 |
| Copper | 0.300 | 0.012 | 0.232 | 0.020 |
| Iron | 0.048 | -0.002 |  | 0.014 |
| Lead | 0.308 | 0.012 | -0.421 | 0.004 |
| Manganese | -0.267 | -0.001 |  |  |
| Molybdenum |  |  | 0.000 | 0.013 |
| Nickel | -0.249 | -0.022 | -0.522 | 0.014 |
| Selenium | 0.384 | -0.020 | 0.343 | 0.008 |
| Silver |  | -0.011 |  |  |
| Strontium | -0.491 | -0.022 |  | 0.002 |
| Thallium | -0.127 | -0.013 | -0.198 | 0.011 |
| Tin | 0.121 | 0.037 | 0.116 | 0.025 |
| Uranium |  |  | 0.477 | 0.007 |
| Vanadium | 0.185 | -0.019 | 0.000 | -0.002 |
| Zinc | 0.252 | -0.009 | 0.439 | 0.016 |

ERS<sup>PR</sup>, ERS plasma risk; ERS<sup>PS</sup>, ERS plasma survival; ERS<sup>UR</sup>, ERS urine risk; ERS<sup>US</sup>, ERS urine survival.

Recognizing the crucial role of gene-environment interactions in ALS pathogenesis, the model was then adjusted for ALS-PGS to account for the cumulative burden of genetic predisposition and environmental exposure. Following adjustment for ALS-PGS (**Supplemental Figure 5; Supplemental Table 7A**), copper, selenium, and zinc were associated with an increased risk of ALS, while antimony and strontium were linked to a decreased risk of ALS. Notably, the ERS^PR^ for metal mixtures remained significantly elevated with OR=2.71 (*p*<0.0001). Next, models with metals and PGS interaction terms were used to exposure the interplay between ALS polygenetic risk, metal exposure, and disease susceptibility. However, only chromium exhibited a statistically significant interaction with the ALS-PGS (*p*<0.01).

Genetic variability in metal metabolism-related can influence how the body processes and responds to metal exposure. Consequently, we incorporated a PGS constructed with genes related to metal metabolism (metal-PGS) into our risk model. Adjustment for metal-PGS reaffirmed associations, with copper, zinc, and selenium linked to increased ALS risk, and antimony, nickel, and strontium with decreased ALS risk (**Supplemental Figure 6; Supplemental Table 7B**). Again, the ERS^PR^ remained significantly elevated, with an OR=2.93 (*p*<0.0001). Moreover, copper and molybdenum showed significant associations with the metal-PGS.

#### Urine

Analysis of urine samples revealed associations between barium, cadmium, copper, molybdenum, selenium, tin, uranium, vanadium, and zinc with increased ALS risk (**Figure 3**); no metals showed associations with decreased risk. Consistent with plasma models, a combination of metals found in urine samples, as captured by the ERS^UR^ (**Table 3; Supplemental Figure 4B**), exhibited a significant association with ALS risk (OR=3.10, *p*>0.001). These findings remained consistent after adjusting for ALS-PGS (**Supplemental Figure 7; Supplemental Table 7C**) and metal-PGS (**Supplemental Figure 8; Supplemental Table 7D**). Significant metal and gene interactions were observed between molybdenum and ALS-PGS, as well as arsenic, barium, nickel, and vanadium with metal-PGS.

**Figure 3.**
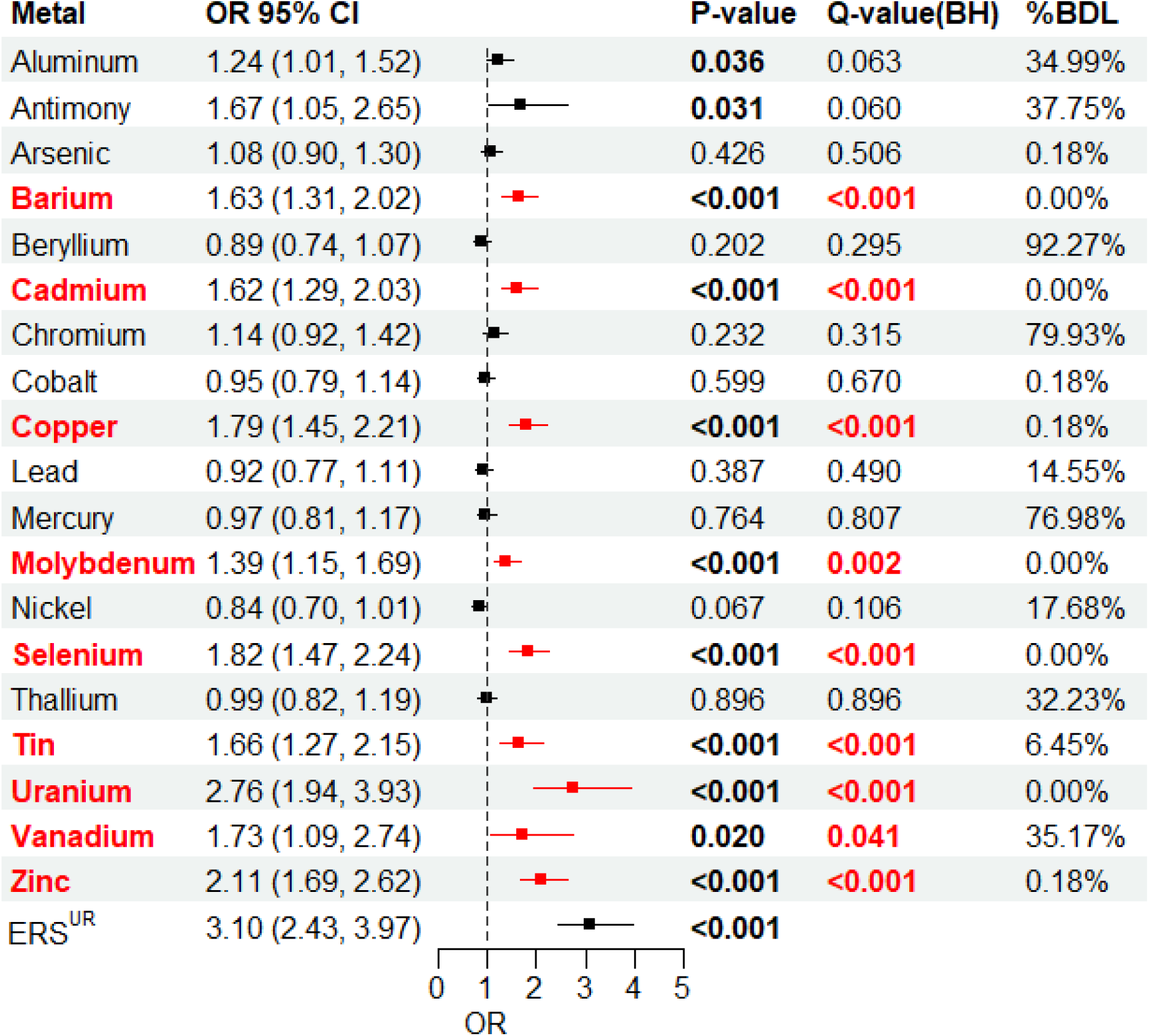
Urine single metal and mixture associations with ALS risk. Single metal logistic regression models based on urine samples where the outcome is case/control status, the predictors are log-transformed, standardized metal levels, and the covariates are continuous age at sample, sex, military service. Mixture metal ERS^UR^ (urine risk environmental risk score). %BDL, the percentage of samples below detection limit; CI, confidence interval; ALS, amyotrophic lateral sclerosis; BH, Benjamini-Hochberg, red font are significant correlations; OR, odds ratio corresponding to one standard deviation increase in log-transformed metals.

### ALS survival

#### Plasma

Only tin exhibited a significant negative individual impact on ALS survival (HR=1.20, *p_adjusted_*<0.05; **Figure 4**). As expected, the ERS^PS^ representing the cumulative effects of all metals included (**Table 3**), was associated with poorer survival (HR=1.42, *p*<0.001; **Supplemental Figure 9A**). Participants with ALS were stratified into quartiles according to their ERS^PS^, revealing a striking difference in survival times between quartile 4 (highest exposure) and quartile 1 (lowest exposure), with individuals in quartile 1 showing an additional survival time of 1.71 years compared to those in quartile 4 (**Figure 5A**). Upon adjustment for ALS-PGS, cadmium, manganese, and tin were found to have a negative impact on ALS survival, with interactions detected between ALS-PGS and aluminum, molybdenum, selenium, and strontium (**Supplemental Figure 10**). Following adjustment for metal-PGS, tin was associated with shorter ALS survival, while iron was linked to improved survival. Significant interactions were identified between metal-PGS and iron, manganese, thallium, and the ERS^PS^ (**Supplemental Figure 11**).

**Figure 4.**
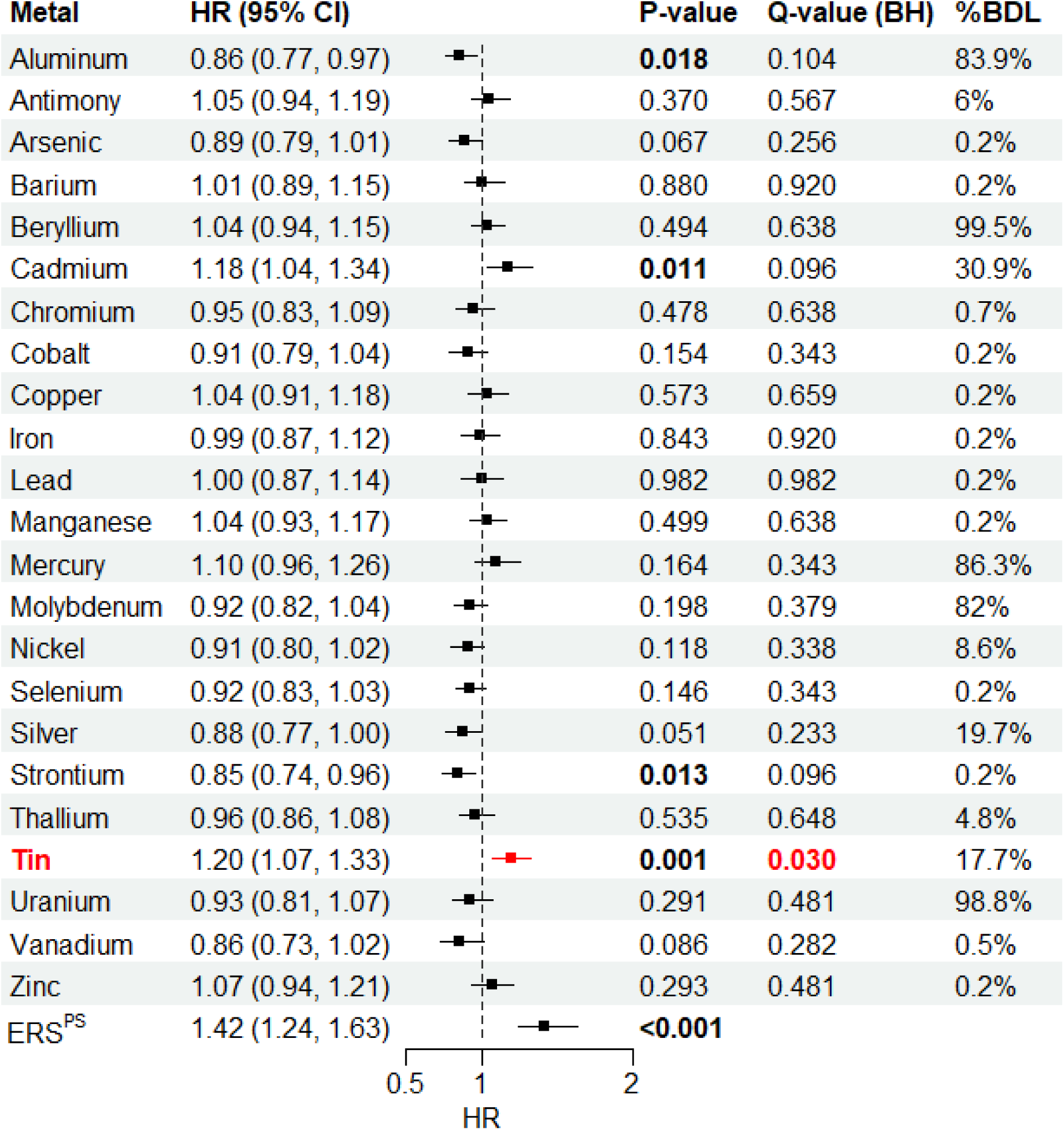
Cohort survival analysis based on plasma samples. Cox proportional hazards survival models from plasma samples with the outcome of survival time since diagnosis (in years), the predictors are log-transformed, standardized metal levels, and adjustment covariates are age at diagnosis, sex, family history of ALS, onset segment, El Escorial criteria, and time between symptom onset and diagnosis. %BDL, the percentage of samples below detection limit; BH, Benjamini-Hochberg, red font are significant correlations; CI, confidence interval; ERS^PS^, plasma survival environmental risk score; HR, hazards ratio corresponding to one standard deviation increase in log-transformed metals.

**Figure 5.**
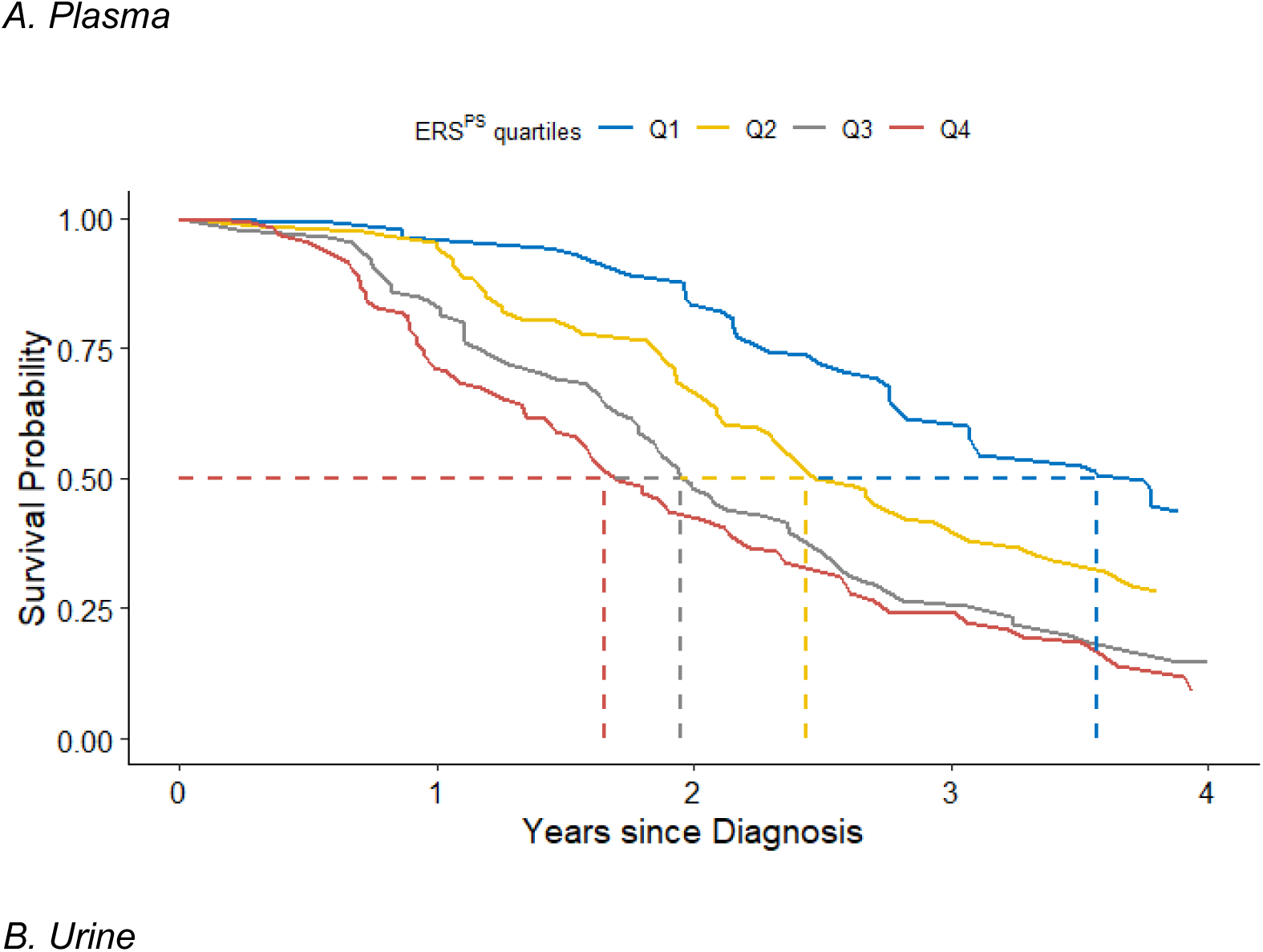
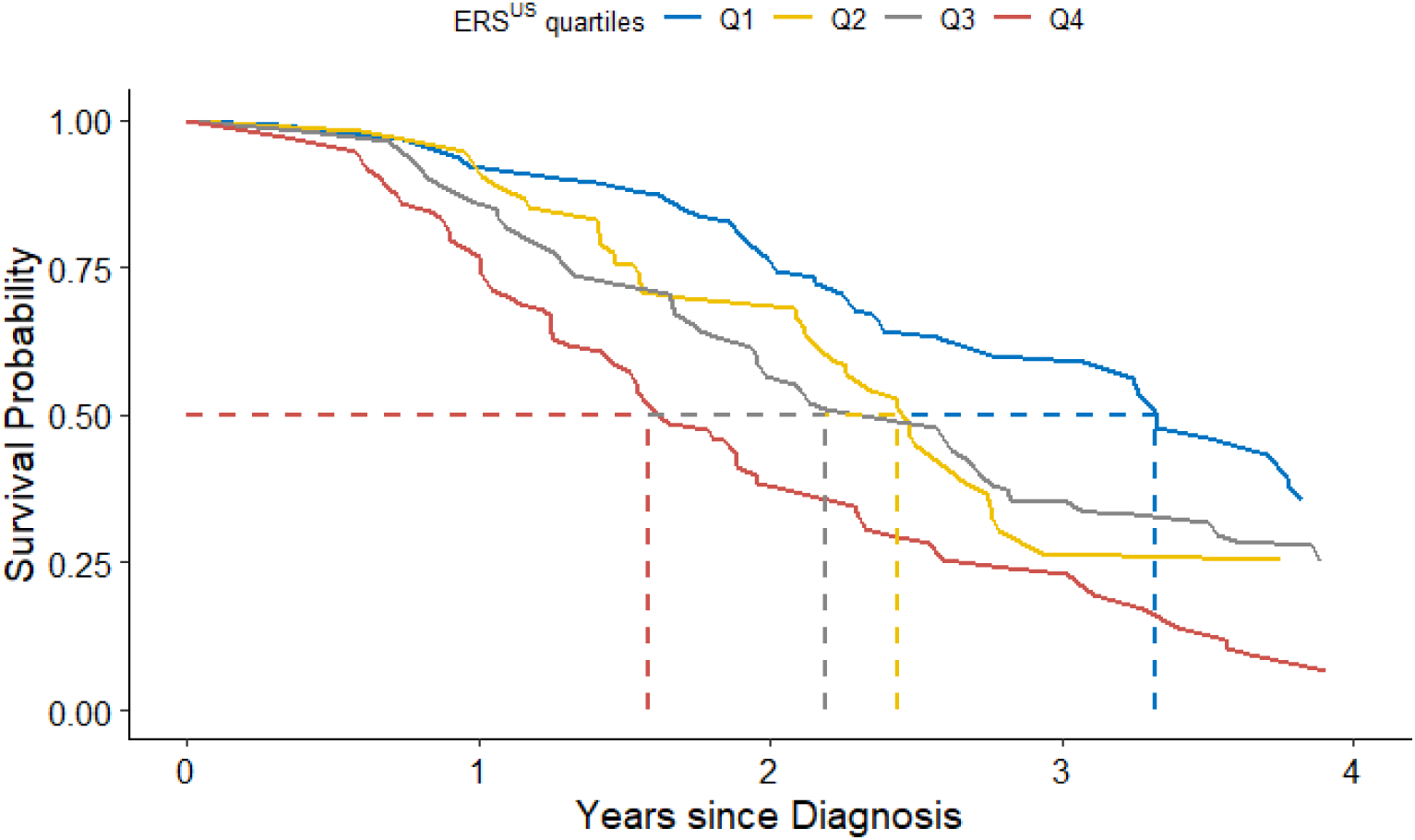
Adjusted survival curves stratified by environmental risk score (ERS) quartile. Kaplan-Meier survival curves of ERS quartiles adjusted for covariates with the inverse probability weights method for (**A**) plasma (ERS^PS)^ and (**B**) urine (ERS^US^) sample sets. Adjusted covariates are age at sample collection, sex, military service, and family history of ALS. Dashed lines indicate the median survival in each ERS strata. The estimated adjusted median survival times for plasma (**A**) are 3.56 years for Quartile 1 (Q1), 2.43 years for Quartile 2 (Q2), 2.35 years for Quartile 3 (Q3), and 1.85 years for Quartile 4 (Q4). Estimated adjusted median survival times for urine (**B**) are 3.25 years for Q1, 2.62 years for Q2, 2.29 years for Q3, and 1.92 years for Q4. Adjusted survival curves and adjusted median survival time estimates are pooled across all 20 imputed datasets.

#### Urine

In contrast to plasma findings, multiple urine metals were associated with shorter ALS survival, including cadmium, copper, nickel, selenium, tin, and zinc (**Figure 6**). However, consistent with plasma results, the ERS^US^ (**Table 3**) once again correlated with shorter ALS survival (HR=1.52, *p*<0.001) (**Supplemental Figure 9B**). Survival curves based on ERS^US^ are depicted in **Figure 5B**, with estimated median survival times of 3.25 years in quartile 1, 2.62 years in quartile 2, 2.29 years in quartile 3, and 1.92 years in quartile 4. This translates to a 1.33-year survival advantage for ALS subjects in quartile 1 compared to those in quartile 4. After adjusting for ALS-PGS, copper, nickel, selenium, tin and zinc continued to correlate with poorer survival, while cadmium did not (**Supplemental Figure 12**). Significant interactions between urine metals and ALS-PGS were detected for copper, lead, nickel, selenium, uranium, and the ERS^US^ (*p_adjusted_*<0.01). No associations were found between metals and ALS survival following adjustment for metal-PGS, although the ERS^US^ remained correlated with shorter survival (**Supplemental Figure 13**). Interactions between metals and metal-PGS were observed for beryllium, copper, tin, vanadium, zinc, and the ERS^US^.

**Figure 6.**
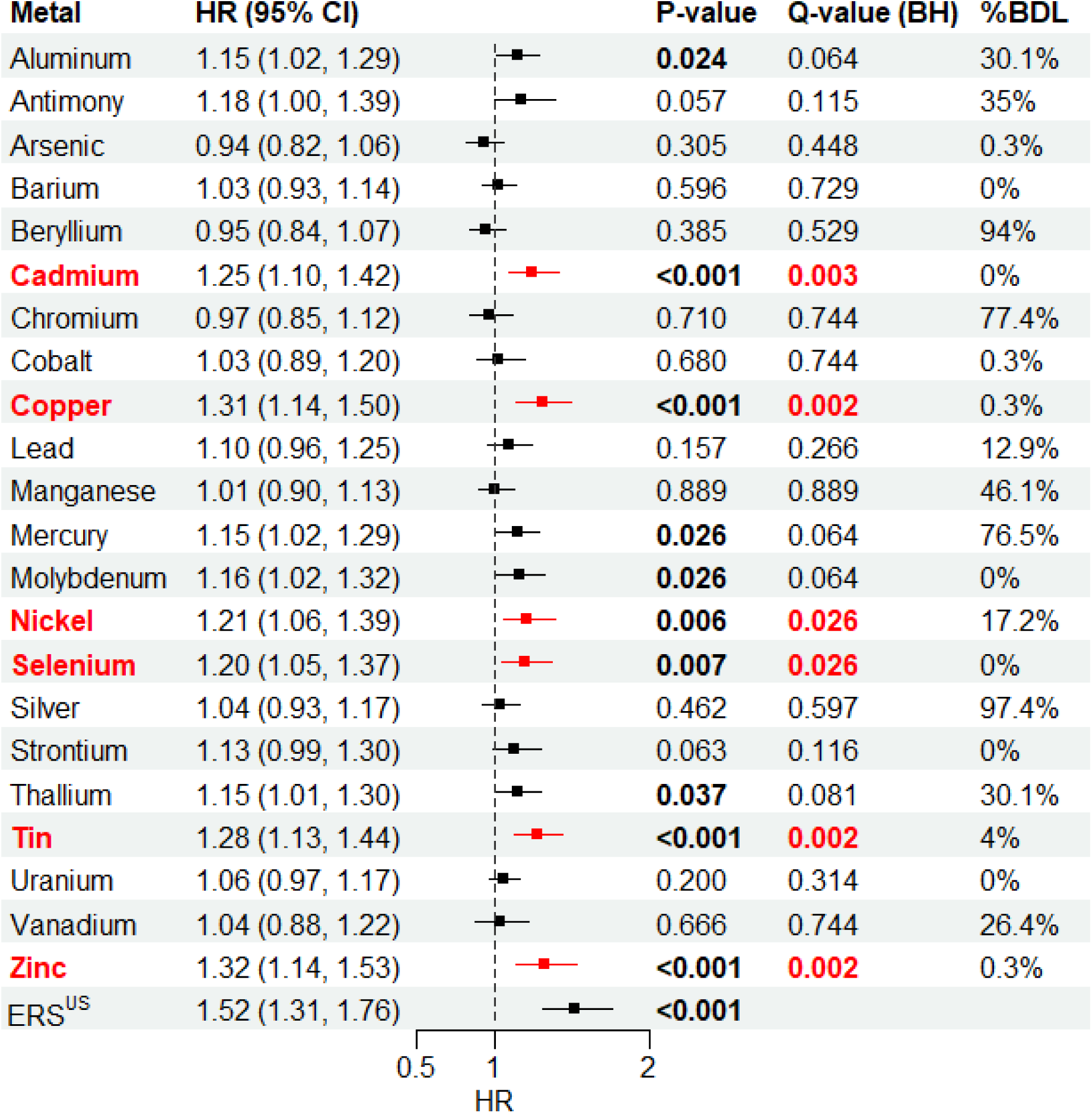
Cohort survival analysis based on urine samples. Cox proportional hazards survival models from urine samples with the outcome of survival time since diagnosis (in years), the predictors are log-transformed, standardized metal levels, and adjustment covariates are age at diagnosis, sex, family history of amyotrophic lateral sclerosis (ALS), onset segment, El Escorial criteria, and time between symptom onset and diagnosis. %BDL, the percentage of samples below detection limit; BH, Benjamini-Hochberg, red font are significant correlations; CI, confidence interval; ERS^US^, urine survival environmental risk score; HR, hazards ratio corresponding to one standard deviation increase in log-transformed metals.

### Self-reported occupations, occupational exposures, non-occupational exposures, and metals levels

Given the associations between self-reported occupational^7^ and non-occupational^21^ metal exposures and ALS risk in this cohort, analyses of occupations, self-reported occupational and non-occupational exposures, and metal levels were conducted.

#### Plasma

ERS^PR^ scores for individuals grouped into an occupational high metal exposure group (job titles of “Building and Grounds Cleaning and Maintenance,” “Construction and Extraction,” “Installation, Maintenance, and Repair,” and “Production Occupations”) were significantly higher than ERS^PR^ scores for those grouped into an occupational low metal exposure group (**Figure 7A**). Furthermore, occupational metal exposures exhibited positive associations with antimony, iron, and thallium levels (**Supplemental Figure 14**). Finally, endorsement of exposure to metals outside the occupational setting positively associated with increased plasma levels of aluminum, cadmium, chromium, lead, manganese, uranium, vanadium, and zinc (**Supplemental Figure 15**).

**Figure 7.**
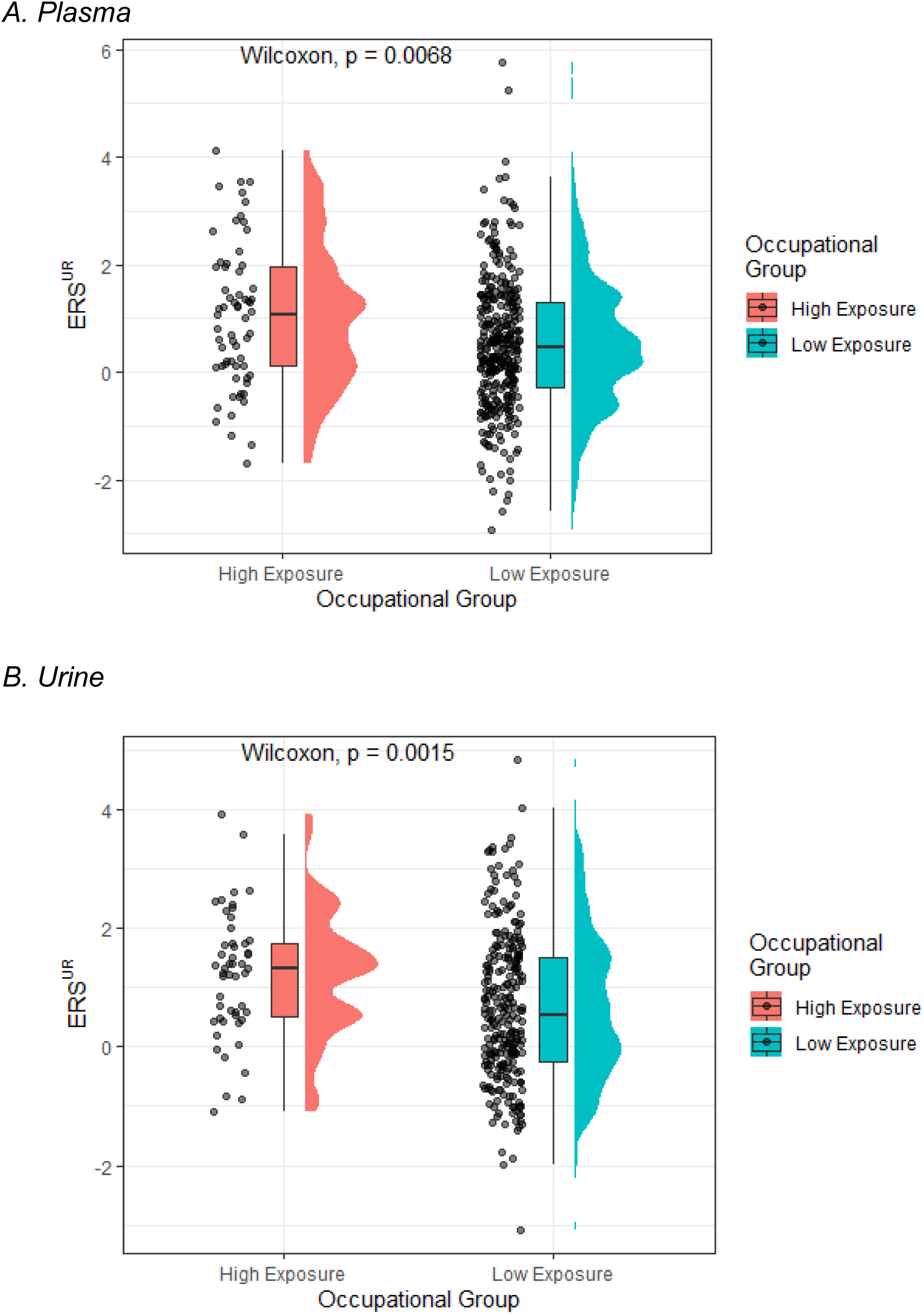
Distribution of environmental risk scores (ERS) by high and low occupational metal exposure. Distribution of the environmental risk scores (ERS) for (**A**) plasma (ERS^PR^) and (**B**) urine (ERS^UR^) by high (plasma, n=99; urine, n=51) and low (plasma, n=292; urine, n=265) occupational metal exposure groups. The high metal exposure group is defined by Standard Occupational Classification (SOC) codes “Building and Grounds Cleaning and Maintenance” (SOC 37), “Construction and Extraction” (SOC 47), “Installation, Maintenance, and Repair” (SOC 49), and “Production Occupations” (SOC 51). All other occupations are included in the low metal exposure group. *P*<0.05 by Wilcoxon test.

#### Urine

Again, individuals stratified into the high metal exposure group based on SOC codes had higher ERS^UR^ score than those in the low metal exposure group (**Figure 7B**). Self-reported occupational metal exposure showed positive associations with mercury and thallium levels (**Supplemental Figure 16**). Additionally, self-reported metal exposure in non-occupational activities demonstrated positive associations with arsenic, cadmium, copper, iron, lead, mercury, molybdenum, nickel, selenium, strontium, thallium, tin, uranium, and zinc (**Supplemental Figure 17**).

These findings underscore the influence of both occupational and recreational activities on metal exposure, highlighting the need for comprehensive assessment of metal exposure sources to understand metal bioaccumulation.

### Supplement usage sensitivity analysis

Finally, recognizing the potential impact of supplement usage on plasma and urine metal levels, a comparison of metal levels characterized by participants’ supplement and/or vitamin usage was performed. In plasma samples, significant differences in median metal concentrations between individuals using supplements and/or vitamins and those abstaining from them were only detected for selenium and vanadium (**Supplemental Table 8**). No significant differences were observed for urine measures (**Supplemental Table 9**). Overall, the use of supplements and/or vitamins does not appear to substantially impact outcomes.

## DISCUSSION

ALS is a complex disease influenced by both genetic and environmental factors. Among these environmental factors, metal exposure is consistently linked to ALS risk.^8^ By measuring metal levels in plasma and urine samples, we confirmed that individual metals, such as copper, selenium, and zinc, are associated with greater ALS risk and shorter survival. Notably, the most robust associations with both risk and survival were observed with cumulative ERS, which represents a mixture of multiple metals. Interestingly, despite the influence of genetic factors on ALS susceptibility, genetic background did not significantly alter the relationship between metal exposure and ALS risk or survival. Finally, we demonstrated that engagement in occupations with heightened metal exposure associates with increased ERS. Overall, our report supports previous research and underscores the pivotal role of metals in both ALS onset and survival within this Michigan-based cohort.

The association between copper and increased ALS risk and decreased survival is consistent with previous clinical studies.^23–25^ Individuals are exposed to copper during their lifetime through various sources, including diet (e.g., seafood, nuts, seeds), drinking water, dietary supplements, cookware, and environmental pollutants. Preclinical models demonstrate that copper accumulation induces oxidative stress,^26, 27^ disrupts mitochondrial function,^28^ and impairs protein folding,^29^ all mechanisms implicated in ALS development. Moreover, clinical data indicate that individuals with ALS often exhibit aberrant copper metabolism, potentially exacerbating neurodegeneration.^30, 31^ While the exact mechanisms linking copper exposure to ALS pathogenesis are not fully understood, accumulating evidence suggests that disturbances in copper levels contribute to development and progression of ALS.

The association between selenium and ALS was first discovered in 1977 through four separate incidences of sporadic ALS among unrelated farmers residing within a 15 km radius of each other in South Dakota,^32^ a region known for selenium-rich soil, which leads to high dietary selenium and selenium intoxication in livestock. Since then, clinical research continues to implicate selenium in ALS pathophysiology.^23, 24, 33–36^ Exposure to selenium occurs through dietary intake of foods, such as seafood, meats, nuts, seeds, and grains grown in selenium-rich soil.^37^ Additionally, selenium exposure can occur through drinking water, dietary supplements, and environmental sources, such as air and soil contamination from industrial activities and agricultural practices. Selenium exposure is believed to influence ALS risk and survival through oxidative stress^38^ and neuroinflammation.^39^ As a component of selenoproteins, such as glutathione peroxidases and thioredoxin reductases, selenium helps neutralize reactive oxygen species, thus mitigating oxidative stress and dampening inflammatory processes.^40^ However, excessive selenium levels cause mitochondrial dysfunction in the cerebral cortex of mice treated with inorganic selenium^41^ and promote SOD1 accumulation in the mitochondria of human neuroblastoma cells treated with selenium,^42^ a feature that is observed in neurons during ALS pathogenesis.^43^ Additionally, selenium exposure-induced neurodegeneration, motor deficiency, and death in *C. elegans* is linked to reduced cholinergic signaling.^44^ Overall, the exact mechanisms by which selenium exposure may affect ALS risk or survival remain unclear and warrant further investigation through both preclinical models and clinical studies.

Similar to the results presented here, previous studies support the association between zinc levels and ALS risk.^23, 25, 45, 46^ Exposure to zinc occurs via dietary sources (e.g., dairy products, nuts, seeds, and whole grains), supplements, environmental pollution (particularly in industrial areas), and consumer products (e.g., cosmetics and sunscreen). As zinc is involved in multiple cellular processes, including synaptic transmission, protein folding, and antioxidant defense mechanisms,^47^ dysregulation of zinc homeostasis can cause excitotoxicity,^48^ impaired protein clearance,^49, 50^ and oxidative stress,^47^ and all of which are implicated in ALS.^51^ Excess zinc also interferes with mitochondrial function^52, 53^ and disrupts calcium homeostasis,^48^ further exacerbating neuronal dysfunction and death. Superoxide dismutase 1 (SOD1) enzyme protects cells from damage caused by oxidative stress^54^ and mutations in the *SOD1* gene are associated with ALS.^54^ Zinc ions are essential for SOD1 stability and enzymatic activity. However, alterations in zinc levels can lead to SOD1 misfolding and aggregation.^55^ Indeed, chronic oral administration of zinc sulfate to transgenic mice overexpressing the human mutated form of SOD1 (G93A) decreases mouse survival.^56^ Additional research is needed to elucidate the role of zinc in ALS pathogenesis and its impact on disease progression and patient outcomes.

In the current study, antimony, nickel, and strontium associated with decreased risk of ALS. Current research investigating the effects of antimony on human health and cellular mechanisms are limited.^57^ However, antimony is detected in human brain^58^ and is associated with neurobehavioral changes and neurotoxicity.^59, 60^ Regarding nickel, a population-based case-control study conducted in Denmark did not detect an association between occupational exposure to nickel and ALS.^61^ It is surprising that nickel would be protective against ALS, considering its ability to accumulate in neuronal tissue, induce apoptosis, inhibit neurotransmission, and promote inflammation.^62, 63^ Similarly, the protective role of strontium conflicts with our previous research examining early-life metal exposure through the analysis of human permanent teeth.^45^ In that study, exposure to strontium during late childhood and adolescence correlated with ALS. This discrepancy could stem from the biosample analyzed, as strontium uptake in teeth tends to occur during the period of tooth formation, reflecting exposure levels during that specific developmental stage.^64^ Thus, the potential protective effects of antimony, nickel, and strontium against ALS warrant further confirmation and thorough investigation.

The use of plasma and urine samples to measure metal exposures offers several advantages, e.g., urine sampling is relatively easy and non-invasive, and both measures reflect integrated exposure. However, measurements must be interpreted cautiously. Levels in plasma and urine may fluctuate due to factors such as diet, hydration status, and circadian rhythms, potentially leading to variability in measurements;^9^ however, it is worth noting that supplement or vitamin use did not impact metal levels in the current study. Additionally, blood and urine often represent more recent, rather than chronic, exposures. For example, while plasma is the biomarker of choice for selenium, its half-life in plasma is only a few days^9^ and plasma levels generally decrease with age.^65^ Moreover, some metals may not be well-represented in plasma or urine. Zinc is primarily distributed intracellularly in the musculoskeletal system and plasma zinc does not represent intracellular accumulation.^9, 66^ Urinary zinc is an accurate biomarker but again represents recent exposures.^67^ Biomarkers such as hair, teeth, and nails can provide a historical record of metal accumulation over time and have been used to offer insights into chronic metal exposure and ALS.^45, 68, 69^ However, challenges remain regarding the suitability of the biomarker, variability in sample quality, and growth rate.^9^ Thus, selecting the appropriate biomarker is crucial for accurately detecting metal levels and assessing exposure.

Most prior epidemiological investigations into the relationship between metals and ALS centered on individual metals, failing to account for the cumulative effect of mixed exposures. Individuals are unlikely to be exposed to a single pollutant over their lifetime; instead, they will encounter multiple pollutants, and an environmental risk score can effectively account for this complexity in exposure. In the current study, we found that mixtures of metals, represented by ERS, had a greater effect on ALS risk and survival than single metal exposures. ERS derived from the quantification of metal levels in biofluids have been previously used to explore the relationship between metal exposure and disease risk and survival across various health domains, including obesity and its comorbidities,^70^ glucose homeostasis,^71^ and heart rate regulation.^72^ Thus, ERS are a valuable tool for studying multi-pollutant exposures and their impact on disease risk and survival.

The gene and environment hypothesis suggests that interactions between genetic factors and environmental exposures play a crucial role in determining an individual’s risk for developing a particular disease.^73^ Several studies have suggested that environmental factors, such as heavy metals, can interact with genetic variants linked to ALS, influencing disease onset, progression, and severity.^74, 75^ Additionally, previous studies have successfully integrated environmental and genetic factors into risk prediction models for cancer.^76, 77^ However, we found that accounting for ALS risk genes or genes involved in metal metabolism did not influence the relationship between metal exposure and ALS. The relationship between genes, environmental exposures (such as metal exposures), and disease risk is often complex and multifactorial. Adjusting for ALS risk genes or genes involved in metal metabolism may not capture the full complexity of these interactions. Additionally, our current knowledge of the genetic basis of ALS and metal metabolism heterogeneity may be incomplete. Other genetic factors or pathways involved in ALS risk or metal metabolism may be unaccounted for in our current PGS.

Finally, in a previous investigation, we explored self-reported occupational exposures and established a significant association between workplace metal exposure and ALS risk.^7^ Moreover, we identified specific SOC codes that were highly likely to report occupational exposure to metals. In the current study, ALS patients were categorized into high or low occupational metal exposure groups based on whether their self-reported occupation fell within SOC codes associated with high exposure, as identified in our previous study. As anticipated, patients in the high metal exposure groups exhibited higher ERS values compared to those in the low metal exposure groups. These results continue to support the finding that occupations linked to metal exposure associate with elevated risk of ALS. Furthermore, occupational and non-occupational exposure^21^ to metals correlated with metal levels detected in plasma and urine samples. This association underscores the potential impact of both occupational and non-occupational activities on overall metal exposure in individuals, emphasizing the necessity of accounting for occupational and environmental sources when evaluating overall exposure risk.

Strengths of this study include its large sample size and the measurement of exposures through biomarkers, thus mitigating recall bias associated with questionnaires. However, there are limitations to consider. First, the study represents a cross-sectional analysis and relying on measurements taken at a single time point fails to capture potential fluctuations in metal exposures and levels over time. Consequently, this snapshot may not accurately reflect lifetime exposures as metal levels are subject to sampling variation and metabolic pathways, potentially altering their concentrations over extended periods. Second, the collection of samples after disease onset may confound results, blurring the distinction between disease progression effects and true baseline measures. Future longitudinal prospective studies will be crucial for tracking changes over time and investigating disease development. While our findings suggest an association between metals and ALS risk and survival, caution is warranted as they do not establish causation. Unlike the ALS-PGS, the metal-PGS does not have a large genome-wide association study from which to draw weights from. Thus, the construction of the metal-PGS is more ad hoc and less robust than standard methods used for the ALS-PGS. Large genetic studies of xenobiotic metabolism would be critical in furthering understanding of gene-environment interactions. Lastly, given the study used a Michigan-based cohort, validating these results across different cohorts is critical to account for geographical variation in metal exposure and ensure broader generalizability.

## Conclusions

To conclude, we demonstrate that metal mixtures, as represented by cumulative ERS, associate with increased ALS risk and reduced ALS survival, regardless of genetic predisposition. These associations correlate with self-reported occupational metal exposure. Utilizing quantitative assessments of metal levels in biofluids like plasma or urine could offer a pathway to predict future ALS risk, providing a promising avenue for early identification and intervention. Overall, these results offer valuable insights into the complexities of ALS pathogenesis, heterogeneity, and progression.

## Acknowledgements

We are indebted to the participants who provided samples. We thank Crystal Pacut, Stacey Jacoby, PhD, Madeleine Batra, Hasan Farid, MS, Caroline Piecuch, Sushant Obeja, and Adam Patterson for study support.

## Data Availability

Sharing of non-identifiable data will be considered at the reasonable request of a qualified investigator.

## REFERENCES

1. Feldman EL, Goutman SA, Petri S, et al. Amyotrophic lateral sclerosis. Lancet 2022.

2. Goutman SA, Savelieff MG, Jang DG, Hur J, Feldman EL. The amyotrophic lateral sclerosis exposome: recent advances and future directions. Nat Rev Neurol 2023;19:617–634.

3. Dou J, Bakulski K, Guo K, et al. Cumulative Genetic Score and C9orf72 Repeat Status Independently Contribute to Amyotrophic Lateral Sclerosis Risk in 2 Case-Control Studies. Neurol Genet 2023;9:e200079.

4. Goutman SA, Boss J, Jang DG, et al. Environmental risk scores of persistent organic pollutants associate with higher ALS risk and shorter survival in a new Michigan case/control cohort. J Neurol Neurosurg Psychiatry 2023.

5. Goutman SA, Boss J, Patterson A, Mukherjee B, Batterman S, Feldman EL. High plasma concentrations of organic pollutants negatively impact survival in amyotrophic lateral sclerosis. J Neurol Neurosurg Psychiatry 2019;90:907–912.

6. Su FC, Goutman SA, Chernyak S, et al. Association of Environmental Toxins With Amyotrophic Lateral Sclerosis. JAMA Neurol 2016;73:803–811.

7. Goutman SA, Boss J, Godwin C, Mukherjee B, Feldman EL, Batterman SA. Associations of self-reported occupational exposures and settings to ALS: a case-control study. Int Arch Occup Environ Health 2022;95:1567–1586.

8. Cicero CE, Mostile G, Vasta R, et al. Metals and neurodegenerative diseases. A systematic review. Environ Res 2017;159:82–94.

9. Martinez-Morata I, Sobel M, Tellez-Plaza M, Navas-Acien A, Howe CG, Sanchez TR. A State-of-the-Science Review on Metal Biomarkers. Curr Environ Health Rep 2023;10:215–249.

10. Goutman SA, Boss J, Iyer G, et al. Body mass index associates with amyotrophic lateral sclerosis survival and metabolomic profiles. Muscle Nerve 2022.

11. Goutman SA, Boss J, Godwin C, Mukherjee B, Feldman EL, Batterman SA. Occupational history associates with ALS survival and onset segment. Amyotroph Lateral Scler Frontotemporal Degener 2022:1–11.

12. Goutman SA, Guo K, Savelieff MG, et al. Metabolomics identifies shared lipid pathways in independent amyotrophic lateral sclerosis cohorts. Brain 2022.

13. Goutman SA, Boss J, Guo K, et al. Untargeted metabolomics yields insight into ALS disease mechanisms. J Neurol Neurosurg Psychiatry 2020;91:1329–1338.

14. Yu Y, Su FC, Callaghan BC, Goutman SA, Batterman SA, Feldman EL. Environmental Risk Factors and Amyotrophic Lateral Sclerosis (ALS): A Case-Control Study of ALS in Michigan. Plos One 2014;9:e101186.

15. van Rheenen W, van der Spek RAA, Bakker MK, et al. Common and rare variant association analyses in amyotrophic lateral sclerosis identify 15 risk loci with distinct genetic architectures and neuron-specific biology. Nat Genet 2021;53:1636–1648.

16. TOXICOLOGICAL PROFILE FOR ARSENIC. Atlanta (GA)2007.

17. DiVincenzo GD, Giordano CJ, Schriever LS. Biologic monitoring of workers exposed to silver. Int Arch Occup Environ Health 1985;56:207–215.

18. Apostoli P, Lucchini R, Alessio L. Are current biomarkers suitable for the assessment of manganese exposure in individual workers? Am J Ind Med 2000;37:283–290.

19. D’Haese PC, Van Landeghem GF, Lamberts LV, Bekaert VA, Schrooten I, De Broe ME. Measurement of strontium in serum, urine, bone, and soft tissues by Zeeman atomic absorption spectrometry. Clin Chem 1997;43:121–128.

20. Du J, Boss J, Han P, et al. Variable selection with multiply-imputed datasets: choosing between stacked and grouped methods. J Comput Graph Stat 2022;31:1063–1075.

21. Goutman SA, Boss J, Jang DG, et al. Avocational exposure associations with ALS risk, survival, and phenotype: A Michigan-based case-control study. J Neurol Sci 2024;457:122899.

22. Rohart F, Gautier B, Singh A, Le Cao KA. mixOmics: An R package for ’omics feature selection and multiple data integration. PLoS Comput Biol 2017;13:e1005752.

23. Roos PM, Vesterberg O, Syversen T, Flaten TP, Nordberg M. Metal concentrations in cerebrospinal fluid and blood plasma from patients with amyotrophic lateral sclerosis. Biol Trace Elem Res 2013;151:159–170.

24. Peters TL, Beard JD, Umbach DM, et al. Blood levels of trace metals and amyotrophic lateral sclerosis. Neurotoxicology 2016;54:119–126.

25. Hozumi I, Hasegawa T, Honda A, et al. Patterns of levels of biological metals in CSF differ among neurodegenerative diseases. J Neurol Sci 2011;303:95–99.

26. Lu Q, Zhang Y, Zhao C, Zhang H, Pu Y, Yin L. Copper induces oxidative stress and apoptosis of hippocampal neuron via pCREB/BDNF/ and Nrf2/HO-1/NQO1 pathway. J Appl Toxicol 2022;42:694–705.

27. Lamtai M, Zghari O, Ouakki S, et al. Chronic copper exposure leads to hippocampus oxidative stress and impaired learning and memory in male and female rats. Toxicol Res 2020;36:359–366.

28. Zischka H, Einer C. Mitochondrial copper homeostasis and its derailment in Wilson disease. Int J Biochem Cell Biol 2018;102:71–75.

29. Zuily L, Lahrach N, Fassler R, et al. Copper Induces Protein Aggregation, a Toxic Process Compensated by Molecular Chaperones. mBio 2022;13:e0325121.

30. Sauzeat L, Bernard E, Perret-Liaudet A, et al. Isotopic Evidence for Disrupted Copper Metabolism in Amyotrophic Lateral Sclerosis. iScience 2018;6:264–271.

31. Barros A, Dourado MET, Jr., Pedrosa LFC, Leite-Lais L. Association of Copper Status with Lipid Profile and Functional Status in Patients with Amyotrophic Lateral Sclerosis. J Nutr Metab 2018;2018:5678698.

32. Kilness AW, Hichberg FH. Amyotrophic lateral sclerosis in a high selenium environment. JAMA 1977;237:2843–2844.

33. Vinceti M, Bottecchi I, Fan A, Finkelstein Y, Mandrioli J. Are environmental exposures to selenium, heavy metals, and pesticides risk factors for amyotrophic lateral sclerosis? Rev Environ Health 2012;27:19–41.

34. Vinceti M, Nacci G, Rocchi E, et al. Mortality in a population with long-term exposure to inorganic selenium via drinking water. J Clin Epidemiol 2000;53:1062–1068.

35. Vinceti M, Solovyev N, Mandrioli J, et al. Cerebrospinal fluid of newly diagnosed amyotrophic lateral sclerosis patients exhibits abnormal levels of selenium species including elevated selenite. Neurotoxicology 2013;38:25–32.

36. Bergomi M, Vinceti M, Nacci G, et al. Environmental exposure to trace elements and risk of amyotrophic lateral sclerosis: a population-based case-control study. Environ Res 2002;89:116–123.

37. Vinceti M, Filippini T, Wise LA. Environmental Selenium and Human Health: an Update. Curr Environ Health Rep 2018;5:464–485.

38. Vinceti M, Mandrioli J, Borella P, Michalke B, Tsatsakis A, Finkelstein Y. Selenium neurotoxicity in humans: bridging laboratory and epidemiologic studies. Toxicol Lett 2014;230:295–303.

39. Naderi M, Puar P, Zonouzi-Marand M, Chivers DP, Niyogi S, Kwong RWM. A comprehensive review on the neuropathophysiology of selenium. Sci Total Environ 2021;767:144329.

40. Cardoso BR, Roberts BR, Bush AI, Hare DJ. Selenium, selenoproteins and neurodegenerative diseases. Metallomics 2015;7:1213–1228.

41. Glaser V, Nazari EM, Muller YM, et al. Effects of inorganic selenium administration in methylmercury-induced neurotoxicity in mouse cerebral cortex. Int J Dev Neurosci 2010;28:631–637.

42. Maraldi T, Riccio M, Zambonin L, Vinceti M, De Pol A, Hakim G. Low levels of selenium compounds are selectively toxic for a human neuron cell line through ROS/RNS increase and apoptotic process activation. Neurotoxicology 2011;32:180–187.

43. Sotelo-Silveira JR, Lepanto P, Elizondo V, et al. Axonal mitochondrial clusters containing mutant SOD1 in transgenic models of ALS. Antioxid Redox Signal 2009;11:1535–1545.

44. Estevez AO, Morgan KL, Szewczyk NJ, Gems D, Estevez M. The neurodegenerative effects of selenium are inhibited by FOXO and PINK1/PTEN regulation of insulin/insulin-like growth factor signaling in Caenorhabditis elegans. Neurotoxicology 2014;41:28–43.

45. Figueroa-Romero C, Mikhail KA, Gennings C, et al. Early life metal dysregulation in amyotrophic lateral sclerosis. Ann Clin Transl Neurol 2020;7:872–882.

46. Pupillo E, Bianchi E, Chio A, et al. Amyotrophic lateral sclerosis and food intake. Amyotroph Lateral Scler Frontotemporal Degener 2018;19:267–274.

47. Chasapis CT, Ntoupa PA, Spiliopoulou CA, Stefanidou ME. Recent aspects of the effects of zinc on human health. Arch Toxicol 2020;94:1443–1460.

48. Inoue K, O’Bryant Z, Xiong ZG. Zinc-permeable ion channels: effects on intracellular zinc dynamics and potential physiological/pathophysiological significance. Curr Med Chem 2015;22:1248–1257.

49. Mezzaroba L, Alfieri DF, Colado Simao AN, Vissoci Reiche EM. The role of zinc, copper, manganese and iron in neurodegenerative diseases. Neurotoxicology 2019;74:230–241.

50. Cristovao JS, Santos R, Gomes CM. Metals and Neuronal Metal Binding Proteins Implicated in Alzheimer’s Disease. Oxid Med Cell Longev 2016;2016:9812178.

51. Feldman EL, Goutman SA, Petri S, et al. Amyotrophic lateral sclerosis. Lancet 2022;400:1363–1380.

52. Gonzalez OA, Novak MJ, Kirakodu S, et al. Effects of aging on apoptosis gene expression in oral mucosal tissues. Apoptosis 2013;18:249–259.

53. Choi S, Liu X, Pan Z. Zinc deficiency and cellular oxidative stress: prognostic implications in cardiovascular diseases. Acta Pharmacol Sin 2018;39:1120–1132.

54. Chattopadhyay M, Valentine JS. Aggregation of copper-zinc superoxide dismutase in familial and sporadic ALS. Antioxid Redox Signal 2009;11:1603–1614.

55. Sannigrahi A, Chowdhury S, Das B, et al. The metal cofactor zinc and interacting membranes modulate SOD1 conformation-aggregation landscape in an in vitro ALS model. Elife 2021;10.

56. Groeneveld GJ, de Leeuw van Weenen J, van Muiswinkel FL, et al. Zinc amplifies mSOD1-mediated toxicity in a transgenic mouse model of amyotrophic lateral sclerosis. Neurosci Lett 2003;352:175–178.

57. Lai Z, He M, Lin C, Ouyang W, Liu X. Interactions of antimony with biomolecules and its effects on human health. Ecotoxicol Environ Saf 2022;233:113317.

58. Hock A, Demmel U, Schicha H, Kasperek K, Feinendegen LE. Trace element concentration in human brain. Activation analysis of cobalt, iron, rubidium, selenium, zinc, chromium, silver, cesium, antimony and scandium. Brain 1975;98:49–64.

59. Tanu T, Anjum A, Jahan M, et al. Antimony-Induced Neurobehavioral and Biochemical Perturbations in Mice. Biol Trace Elem Res 2018;186:199–207.

60. Wang X, Zhu P, Xu S, et al. Antimony, a novel nerve poison, triggers neuronal autophagic death via reactive oxygen species-mediated inhibition of the protein kinase B/mammalian target of rapamycin pathway. Int J Biochem Cell Biol 2019;114:105561.

61. Dickerson AS, Hansen J, Gredal O, Weisskopf MG. Study of Occupational Chromium, Iron, and Nickel Exposure and Amyotrophic Lateral Sclerosis in Denmark. Int J Environ Res Public Health 2020;17.

62. Song X, Fiati Kenston SS, Kong L, Zhao J. Molecular mechanisms of nickel induced neurotoxicity and chemoprevention. Toxicology 2017;392:47–54.

63. Saito M, Arakaki R, Yamada A, Tsunematsu T, Kudo Y, Ishimaru N. Molecular Mechanisms of Nickel Allergy. Int J Mol Sci 2016;17.

64. Reiss LZ. Strontium-90 absorption by deciduous teeth. Science 1961;134:1669–1673.

65. Maehira F, Luyo GA, Miyagi I, et al. Alterations of serum selenium concentrations in the acute phase of pathological conditions. Clin Chim Acta 2002;316:137–146.

66. Lowe NM, Fekete K, Decsi T. Methods of assessment of zinc status in humans: a systematic review. Am J Clin Nutr 2009;89:2040S–2051S.

67. Taylor A. Detection and monitoring of disorders of essential trace elements. Ann Clin Biochem 1996;33 ( Pt 6):486–510.

68. Andrew AS, Chen CY, Caller TA, et al. Toenail mercury Levels are associated with amyotrophic lateral sclerosis risk. Muscle Nerve 2018.

69. Andrew AS, O’Brien KM, Jackson BP, et al. Keratinous biomarker of mercury exposure associated with amyotrophic lateral sclerosis risk in a nationwide U.S. study. Amyotroph Lateral Scler Frontotemporal Degener 2020;21:420–427.

70. Wang X, Mukherjee B, Park SK. Associations of cumulative exposure to heavy metal mixtures with obesity and its comorbidities among U.S. adults in NHANES 2003-2014. Environ Int 2018;121:683–694.

71. Wang X, Mukherjee B, Karvonen-Gutierrez CA, et al. Urinary metal mixtures and longitudinal changes in glucose homeostasis: The Study of Women’s Health Across the Nation (SWAN). Environ Int 2020;145:106109.

72. Fu Y, Liu Y, Liu Y, et al. Relationship between cumulative exposure to metal mixtures and heart rate among Chinese preschoolers. Chemosphere 2022;300:134548.

73. Goutman SA, Hardiman O, Al-Chalabi A, et al. Emerging insights into the complex genetics and pathophysiology of amyotrophic lateral sclerosis. Lancet Neurol 2022;21:465–479.

74. Bailey JM, Colon-Rodriguez A, Atchison WD. Evaluating a Gene-Environment Interaction in Amyotrophic Lateral Sclerosis: Methylmercury Exposure and Mutated SOD1. Curr Environ Health Rep 2017;4:200–207.

75. Morahan JM, Yu B, Trent RJ, Pamphlett R. Genetic susceptibility to environmental toxicants in ALS. Am J Med Genet B Neuropsychiatr Genet 2007;144B:885–890.

76. Archambault AN, Jeon J, Lin Y, et al. Risk Stratification for Early-Onset Colorectal Cancer Using a Combination of Genetic and Environmental Risk Scores: An International Multi-Center Study. J Natl Cancer Inst 2022;114:528–539.

77. Guan Z, Raut JR, Weigl K, et al. Individual and joint performance of DNA methylation profiles, genetic risk score and environmental risk scores for predicting breast cancer risk. Mol Oncol 2020;14:42–53.

