## Supplemental Tables and Figures for "Metal mixtures associate with higher amyotrophic lateral sclerosis risk and mortality independent of genetic risk and correlate to self-reported exposures: a case-control study"

#### Supplemental Table 1. Single nucleotide polymorphisms (SNPs) included in the amyotrophic lateral sclerosis-polygenic risk score (ALS-PGS).

Table contains the identifier, alleles, weight, and *p*-value for each SNP included in the ALS-PGS. This table is provided as a separate file labeled “Supplemental Table 1 – ALS-PGS SNPs.csv.”

#### Supplemental Table 2. Single nucleotide polymorphisms (SNPs) included in the metal-polygenic risk score (metal-PGS).

Table contains the identifier, alleles, chromosome, base pair, and reference for each SNP included in the metal-PGS. The SNPs are based on a literature search. This table is provided as a separate file labeled “Supplemental Table 2 – Metal-PGS SNPs.csv.”

#### Supplemental Table 3. Standard occupational classification (SOC) codes

| **SOC Code** | **Occupation** |
| --- | --- |
| 11-0000 | Management Occupations |
| 13-0000 | Business and Financial Operations Occupations |
| 15-0000 | Computer and Mathematical Occupations |
| 17-0000 | Architecture and Engineering Occupations |
| 19-0000 | Life, Physical, and Social Science Occupations |
| 21-0000 | Community and Social Services Occupations |
| 23-0000 | Legal Occupations |
| 25-0000 | Education, Training, and Library Occupations |
| 27-0000 | Arts, Design, Entertainment, Sports, and Media Occupations |
| 29-0000 | Healthcare Practitioners and Technical Occupations |
| 31-0000 | Healthcare Support Occupations |
| 33-0000 | Protective Service Occupations |
| 35-0000 | Food Preparation and Serving Related Occupations |
| 37-0000 | Building and Grounds Cleaning and Maintenance Occupations |
| 39-0000 | Personal Care and Service Occupations |
| 41-0000 | Sales and Related Occupations |
| 43-0000 | Office and Administrative Support Occupations |
| 45-0000 | Farming, Fishing, and Forestry Occupations |
| 47-0000 | Construction and Extraction Occupations |
| 49-0000 | Installation, Maintenance, and Repair Occupations |
| 52-0000 | Production Occupations |
| 53-0000 | Transportation and Material Moving Occupations |
| 55-0000 | Military Occupations |

#### Supplemental Table 4. ALS and control participant demographics: Plasma and urine

| **Covariate** | **ALS**, N = 255^1^ | **Control**, N = 181^1^ | **p-value**^2^ |
| --- | --- | --- | --- |
| Age at Urine Sample Collection | 65 (58-72) | 61 (55-68) | **<0.001** |
| Age at Plasma Sample Collection | 66 (58-72) | 61 (55-68) | **<0.001** |
| Sex |  |  | **<0.001** |
| Female | 99 (39%) | 101 (56%) |  |
| Male | 156 (61%) | 80 (44%) |  |
| Ethnicity |  |  | **0.018** |
| Hispanic or Latino | 2 (0.8%) | 8 (4.6%) |  |
| Not Hispanic or Latino | 251 (99%) | 166 (95%) |  |
| Missing | 2 | 7 |  |
| Race |  |  | 0.2 |
| Asian | 0 (0%) | 2 (1.1%) |  |
| Black or African American | 8 (3.1%) | 9 (5.0%) |  |
| Other | 1 (0.4%) | 0 (0%) |  |
| White or Caucasian | 245 (96%) | 170 (94%) |  |
| Missing | 1 | 0 |  |
| Military Service |  |  | 0.5 |
| Yes | 30 (12%) | 16 (10%) |  |
| No | 217 (88%) | 142 (90%) |  |
| Missing | 8 | 23 |  |
| Education |  |  | **<0.001** |
| High school or less | 62 (25%) | 8 (5.0%) |  |
| Bachelor’s degree | 56 (23%) | 64 (40%) |  |
| Some postsecondary | 88 (35%) | 29 (18%) |  |
| Graduate degree | 42 (17%) | 58 (36%) |  |
| Missing | 7 | 22 |  |
| Family History of ALS | 26 (10%) | 0 (0%) | **<0.001** |
| Missing | 6 |  |  |
| Age at Diagnosis | 65 (58-71) |  |  |
| Missing | 2 |  |  |
| Diagnosis |  |  |  |
| ALS | 233 (91%) |  |  |
| ALS/Frontotemporal dementia | 13 (5.1%) |  |  |
| Brachial amyotrophic diplegia | 5 (2.0%) |  |  |
| Flail leg | 1 (0.4%) |  |  |
| Progressive muscular atrophy | 3 (1.2%) |  |  |
| El Escorial Criteria |  |  |  |
| Definite | 72 (28%) |  |  |
| Probable | 73 (29%) |  |  |
| Probable, lab supported | 70 (28%) |  |  |
| Possible | 26 (10%) |  |  |
| Suspected | 12 (4.7%) |  |  |
| Missing | 2 |  |  |
| Onset Segment |  |  |  |
| Bulbar | 68 (27%) |  |  |
| Cervical | 90 (36%) |  |  |
| Lumbar | 90 (36%) |  |  |
| Respiratory | 3 (1.2%) |  |  |
| Thoracic | 2 (0.8%) |  |  |
| Missing | 2 |  |  |
| ALSFRS-R | 38 (34-41) |  |  |
| Missing | 1 |  |  |
| Time Between Symptom Onset and Diagnosis | 1.05 (0.61-1.79) |  |  |
| Missing | 4 |  |  |
| Time Between Diagnosis and Urine Sample (Years) | 0.45 (0.21-0.82) |  |  |
| Missing | 2 |  |  |
| Time Between Diagnosis and Plasma Sample (Years) | 0.50 (0.21-0.96) |  |  |
| Missing | 2 |  |  |
| ^1^Median (25%-75%); n (%) | | | |
| ^2^Wilcoxon rank sum test; Pearson’s Chi-squared test; Fisher’s exact test | | | |
| Abbreviations: ALS, amyotrophic lateral sclerosis; ALSFRS-R, Revised ALS Functional Rating Scale | | | |

#### Supplementary Table 5. Metal levels in plasma samples

Abbreviations: ALS, amyotrophic lateral sclerosis; N, the number of subjects in the group; SD, standard deviation; Q10, 10^th^ Percentile; Q25, 25^th^ Percentile; Q50, median; Q75, 75^th^ Percentile; Q90, 90^th^ Percentile. P-value, the *p*-value from the Wilcoxon rank sum test compares metal levels between the case and control groups; BH, Benjamini-Hochberg; %BDL, the percentage of samples below the detection limit. Red, median value is higher in ALS group; Blue, median value is higher in control group; Significance based on *q*-value.

|  | | **ALS, N=387  (μg/ℓ)** | | | | | | | | | **Control, N=281  (μg/ℓ)** | | | | | | | | |  | |
| --- | --- | --- | --- | --- | --- | --- | --- | --- | --- | --- | --- | --- | --- | --- | --- | --- | --- | --- | --- | --- | --- |
| **Metal** | **%BDL** | **Mean** | **SD** | **Min** | **Q10** | **Q25** | **Q50** | **Q75** | **Q90** | **Max** | **Mean** | **SD** | **Min** | **Q10** | **Q25** | **Q50** | **Q75** | **Q90** | **Max** | **P-value** | **Q-value (BH)** |
| Aluminum | 84.0% | 49.56 | 320.11 | 14.14 | 14.14 | 14.14 | 14.14 | 14.14 | 25.75 | 4,612.83 | 64.43 | 587.46 | 14.14 | 14.14 | 14.14 | 14.14 | 14.14 | 23.04 | 9,595.32 | 0.864 | >0.999 |
| Antimony | 3.5% | 3.16 | 2.02 | 0.08 | 1.76 | 2.43 | 3.09 | 3.73 | 4.45 | 23.36 | 3.67 | 1.94 | 1.68 | 2.30 | 2.68 | 3.24 | 4.07 | 5.62 | 28.02 | <0.001 | 0.003 |
| Arsenic | 0.1% | 0.52 | 0.92 | 0.09 | 0.15 | 0.18 | 0.25 | 0.42 | 1.08 | 8.11 | 0.70 | 1.90 | 0.12 | 0.19 | 0.23 | 0.31 | 0.57 | 1.27 | 29.29 | <0.001 | <0.001 |
| Barium | 0.1% | 1.62 | 1.32 | 0.22 | 0.58 | 0.82 | 1.33 | 2.05 | 2.89 | 15.42 | 2.46 | 6.09 | 0.22 | 0.51 | 0.94 | 1.64 | 2.51 | 4.08 | 95.58 | 0.001 | 0.018 |
| Beryllium | 99.6% | 0.07 | 0.00 | 0.07 | 0.07 | 0.07 | 0.07 | 0.07 | 0.07 | 0.11 | 0.07 | 0.02 | 0.07 | 0.07 | 0.07 | 0.07 | 0.07 | 0.07 | 0.34 | 0.763 | >0.999 |
| Cadmium | 36.3% | 0.01 | 0.02 | 0.00 | 0.00 | 0.00 | 0.01 | 0.02 | 0.03 | 0.09 | 0.01 | 0.02 | 0.00 | 0.00 | 0.00 | 0.01 | 0.01 | 0.02 | 0.25 | <0.001 | 0.016 |
| Chromium | 0.4% | 0.65 | 2.15 | 0.06 | 0.23 | 0.28 | 0.37 | 0.49 | 0.82 | 39.08 | 0.53 | 1.00 | 0.09 | 0.24 | 0.29 | 0.35 | 0.47 | 0.62 | 11.65 | 0.543 | >0.999 |
| Cobalt | 0.1% | 0.36 | 1.04 | 0.09 | 0.14 | 0.16 | 0.19 | 0.26 | 0.37 | 13.40 | 0.29 | 0.33 | 0.09 | 0.14 | 0.17 | 0.20 | 0.28 | 0.45 | 3.89 | 0.135 | >0.999 |
| Copper | 0.1% | 1,112.83 | 205.24 | 587.71 | 878.43 | 968.38 | 1,088.97 | 1,231.59 | 1,384.44 | 1,894.29 | 1,065.57 | 226.63 | 671.57 | 822.01 | 921.63 | 1,038.48 | 1,181.87 | 1,326.67 | 2,432.46 | 0.001 | 0.018 |
| Iron | 0.1% | 1,307.97 | 819.60 | 237.89 | 751.77 | 923.79 | 1,149.08 | 1,439.91 | 1,866.88 | 9,084.29 | 1,335.07 | 635.83 | 448.96 | 744.54 | 932.36 | 1,234.62 | 1,569.71 | 2,061.99 | 6,207.42 | 0.130 | >0.999 |
| Lead | 0.8% | 0.46 | 0.57 | 0.03 | 0.10 | 0.18 | 0.29 | 0.49 | 0.94 | 4.09 | 0.37 | 1.12 | 0.02 | 0.06 | 0.11 | 0.16 | 0.26 | 0.55 | 16.40 | <0.001 | <0.001 |
| Manganese | 0.1% | 1.45 | 4.15 | 0.24 | 0.38 | 0.47 | 0.55 | 0.72 | 3.27 | 70.17 | 1.19 | 2.21 | 0.29 | 0.42 | 0.51 | 0.60 | 0.74 | 1.27 | 21.91 | 0.010 | 0.119 |
| Mercury | 85.2% | 1.20 | 1.19 | 0.88 | 0.88 | 0.88 | 0.88 | 0.88 | 1.53 | 11.46 | 1.25 | 1.43 | 0.88 | 0.88 | 0.88 | 0.88 | 0.88 | 2.07 | 16.50 | 0.299 | >0.999 |
| Molybdenum | 85.0% | 2.15 | 1.04 | 1.75 | 1.75 | 1.75 | 1.75 | 1.75 | 3.17 | 8.11 | 2.01 | 0.99 | 1.75 | 1.75 | 1.75 | 1.75 | 1.75 | 2.61 | 12.01 | 0.019 | 0.211 |
| Nickel | 7.7% | 0.82 | 0.63 | 0.20 | 0.33 | 0.51 | 0.76 | 0.97 | 1.24 | 8.56 | 0.97 | 1.23 | 0.20 | 0.37 | 0.59 | 0.85 | 1.06 | 1.30 | 17.28 | 0.006 | 0.080 |
| Selenium | 0.1% | 142.50 | 29.51 | 70.80 | 116.59 | 126.96 | 139.18 | 150.39 | 171.80 | 361.30 | 132.82 | 15.61 | 90.07 | 114.41 | 122.38 | 132.27 | 141.86 | 150.45 | 225.49 | <0.001 | <0.001 |
| Silver | 17.9% | 0.15 | 0.32 | 0.02 | 0.02 | 0.04 | 0.08 | 0.16 | 0.30 | 5.37 | 0.12 | 0.16 | 0.02 | 0.02 | 0.04 | 0.07 | 0.14 | 0.26 | 1.59 | 0.830 | >0.999 |
| Strontium | 0.1% | 25.03 | 13.71 | 8.14 | 14.41 | 17.71 | 22.54 | 28.67 | 36.77 | 203.01 | 30.17 | 18.71 | 11.79 | 17.11 | 22.45 | 26.85 | 34.37 | 42.19 | 254.24 | <0.001 | <0.001 |
| Thallium | 2.8% | 0.03 | 0.01 | 0.01 | 0.01 | 0.02 | 0.03 | 0.04 | 0.05 | 0.11 | 0.03 | 0.01 | 0.01 | 0.02 | 0.02 | 0.03 | 0.04 | 0.05 | 0.09 | 0.031 | 0.309 |
| Tin | 19.3% | 0.39 | 0.97 | 0.07 | 0.07 | 0.12 | 0.19 | 0.35 | 0.66 | 15.05 | 0.27 | 0.32 | 0.07 | 0.07 | 0.11 | 0.18 | 0.30 | 0.53 | 3.72 | 0.166 | >0.999 |
| Uranium | 98.6% | 0.02 | 0.01 | 0.02 | 0.02 | 0.02 | 0.02 | 0.02 | 0.02 | 0.18 | 0.02 | 0.02 | 0.02 | 0.02 | 0.02 | 0.02 | 0.02 | 0.02 | 0.42 | 0.618 | >0.999 |
| Vanadium | 0.3% | 0.12 | 0.48 | 0.01 | 0.03 | 0.04 | 0.05 | 0.07 | 0.14 | 8.14 | 0.10 | 0.54 | 0.02 | 0.03 | 0.03 | 0.04 | 0.06 | 0.08 | 8.36 | <0.001 | <0.001 |
| Zinc | 0.1% | 829.76 | 198.28 | 429.59 | 635.10 | 704.72 | 790.08 | 916.18 | 1,067.51 | 1,738.72 | 771.77 | 181.35 | 507.28 | 612.23 | 661.45 | 740.94 | 837.65 | 943.61 | 2,217.53 | <0.001 | <0.001 |

#### Supplementary Table 6. Metal levels in urine samples

Abbreviations: ALS, amyotrophic lateral sclerosis; N, the number of subjects in the group; SD, standard deviation; Q10, 10^th^ Percentile; Q25, 25^th^ Percentile; Q50, median; Q75, 75^th^ Percentile; Q90, 90^th^ Percentile. P-value, the p-value from the Wilcoxon rank sum test compares metal levels between the case and control groups.; BH, Benjamini-Hochberg; %BDL, the percentage of samples below the detection limit. Red, median value is higher in ALS group; Significance based on *q*-value.

|  | | **ALS, N=322  (μg/ℓ)** | | | | | | | | | **Control, N=194  (μg/ℓ)** | | | | | | | | |  | |
| --- | --- | --- | --- | --- | --- | --- | --- | --- | --- | --- | --- | --- | --- | --- | --- | --- | --- | --- | --- | --- | --- |
| **Metal** | **%BDL** | **Mean** | **SD** | **Min** | **Q10** | **Q25** | **Q50** | **Q75** | **Q90** | **Max** | **Mean** | **SD** | **Min** | **Q10** | **Q25** | **Q50** | **Q75** | **Q90** | **Max** | **P-value** | **Q-value (BH)** |
| Aluminum | 35.0% | 14.58 | 36.52 | 2.84 | 2.84 | 2.84 | 5.96 | 10.91 | 22.65 | 364.19 | 12.87 | 33.96 | 2.84 | 2.84 | 2.84 | 4.51 | 9.20 | 19.13 | 306.75 | 0.003 | 0.031 |
| Antimony | 37.8% | 0.08 | 0.49 | 0.01 | 0.01 | 0.01 | 0.03 | 0.06 | 0.11 | 8.73 | 0.04 | 0.05 | 0.01 | 0.01 | 0.01 | 0.02 | 0.05 | 0.08 | 0.43 | 0.015 | 0.150 |
| Arsenic | 0.2% | 16.92 | 30.56 | 0.22 | 2.38 | 3.77 | 7.64 | 15.17 | 33.85 | 239.70 | 24.16 | 159.09 | 1.15 | 2.13 | 3.86 | 6.82 | 12.92 | 29.81 | 2,212.14 | 0.240 | >0.999 |
| Barium | 0.0% | 3.49 | 13.68 | 0.11 | 0.46 | 0.90 | 1.83 | 3.12 | 6.65 | 242.05 | 1.93 | 3.05 | 0.04 | 0.29 | 0.56 | 1.10 | 2.22 | 3.81 | 32.75 | <0.001 | <0.001 |
| Beryllium | 92.3% | 0.02 | 0.00 | 0.02 | 0.02 | 0.02 | 0.02 | 0.02 | 0.02 | 0.06 | 0.02 | 0.01 | 0.02 | 0.02 | 0.02 | 0.02 | 0.02 | 0.03 | 0.07 | 0.063 | 0.507 |
| Cadmium | 0.0% | 0.49 | 0.53 | 0.02 | 0.11 | 0.18 | 0.33 | 0.58 | 0.98 | 3.77 | 0.31 | 0.32 | 0.01 | 0.05 | 0.12 | 0.22 | 0.38 | 0.65 | 1.76 | <0.001 | <0.001 |
| Chromium | 79.9% | 0.94 | 1.62 | 0.53 | 0.53 | 0.53 | 0.53 | 0.53 | 1.26 | 18.40 | 0.77 | 1.37 | 0.53 | 0.53 | 0.53 | 0.53 | 0.53 | 0.98 | 17.91 | 0.056 | 0.507 |
| Cobalt | 0.2% | 0.63 | 2.68 | 0.01 | 0.07 | 0.11 | 0.18 | 0.30 | 0.55 | 36.65 | 0.43 | 0.80 | 0.01 | 0.06 | 0.09 | 0.20 | 0.38 | 0.91 | 8.41 | 0.314 | >0.999 |
| Copper | 0.2% | 13.56 | 22.40 | 0.33 | 3.69 | 6.40 | 9.90 | 15.18 | 22.25 | 277.36 | 8.49 | 9.97 | 0.68 | 1.79 | 3.50 | 6.55 | 10.82 | 15.91 | 96.77 | <0.001 | <0.001 |
| Iron | 26.3% | 17.79 | 31.09 | 3.07 | 3.07 | 6.09 | 10.19 | 17.38 | 34.14 | 382.91 | 8.14 | 16.77 | 3.07 | 3.07 | 3.07 | 4.99 | 8.53 | 13.37 | 179.67 | <0.001 | <0.001 |
| Lead | 14.5% | 0.19 | 0.12 | 0.04 | 0.04 | 0.10 | 0.17 | 0.24 | 0.34 | 0.85 | 0.20 | 0.16 | 0.04 | 0.04 | 0.08 | 0.17 | 0.27 | 0.38 | 1.08 | 0.899 | >0.999 |
| Manganese | 49.9% | 0.21 | 0.42 | 0.06 | 0.06 | 0.06 | 0.09 | 0.16 | 0.38 | 3.89 | 0.16 | 0.31 | 0.06 | 0.06 | 0.06 | 0.06 | 0.13 | 0.24 | 3.15 | 0.006 | 0.068 |
| Mercury | 77.0% | 0.39 | 0.27 | 0.28 | 0.28 | 0.28 | 0.28 | 0.28 | 0.68 | 2.76 | 0.39 | 0.26 | 0.28 | 0.28 | 0.28 | 0.28 | 0.28 | 0.73 | 1.93 | 0.959 | >0.999 |
| Molybdenum | 0.0% | 59.32 | 54.17 | 1.97 | 13.84 | 25.58 | 43.68 | 74.14 | 123.62 | 423.66 | 47.32 | 53.18 | 1.96 | 8.05 | 13.87 | 30.17 | 56.09 | 109.45 | 331.18 | <0.001 | <0.001 |
| Nickel | 17.7% | 1.47 | 1.59 | 0.18 | 0.18 | 0.47 | 1.12 | 1.93 | 3.05 | 15.04 | 1.84 | 2.39 | 0.18 | 0.18 | 0.37 | 1.03 | 2.05 | 4.35 | 14.75 | 0.809 | >0.999 |
| Selenium | 0.0% | 54.91 | 45.33 | 2.95 | 15.02 | 26.97 | 42.29 | 69.09 | 104.98 | 353.37 | 35.03 | 30.67 | 2.52 | 7.45 | 13.26 | 28.98 | 46.81 | 69.93 | 259.40 | <0.001 | <0.001 |
| Silver | 98.0% | 0.02 | 0.08 | 0.02 | 0.02 | 0.02 | 0.02 | 0.02 | 0.02 | 1.51 | 0.02 | 0.00 | 0.02 | 0.02 | 0.02 | 0.02 | 0.02 | 0.02 | 0.04 | 0.178 | >0.999 |
| Strontium | 0.0% | 158.38 | 134.84 | 8.70 | 48.48 | 78.58 | 126.17 | 195.99 | 283.23 | 1,351.11 | 110.18 | 102.53 | 5.48 | 25.49 | 43.12 | 88.00 | 148.27 | 217.34 | 922.83 | <0.001 | <0.001 |
| Thallium | 32.2% | 0.27 | 0.32 | 0.04 | 0.04 | 0.04 | 0.16 | 0.34 | 0.64 | 2.65 | 0.26 | 0.32 | 0.04 | 0.04 | 0.04 | 0.14 | 0.33 | 0.71 | 1.98 | 0.403 | >0.999 |
| Tin | 6.4% | 1.81 | 9.03 | 0.02 | 0.10 | 0.20 | 0.42 | 1.16 | 2.59 | 149.69 | 0.60 | 1.06 | 0.02 | 0.02 | 0.10 | 0.28 | 0.64 | 1.28 | 9.68 | <0.001 | <0.001 |
| Uranium | 0.0% | 1.30 | 8.86 | 0.04 | 0.17 | 0.30 | 0.52 | 0.88 | 1.34 | 152.48 | 0.38 | 0.35 | 0.02 | 0.09 | 0.15 | 0.31 | 0.49 | 0.75 | 2.81 | <0.001 | <0.001 |
| Vanadium | 35.2% | 0.16 | 0.96 | 0.02 | 0.02 | 0.02 | 0.06 | 0.12 | 0.20 | 16.88 | 0.07 | 0.14 | 0.02 | 0.02 | 0.02 | 0.02 | 0.07 | 0.11 | 1.61 | <0.001 | <0.001 |
| Zinc | 0.2% | 655.58 | 756.37 | 5.94 | 135.59 | 250.62 | 470.49 | 812.11 | 1,213.73 | 6,717.82 | 323.94 | 324.62 | 10.83 | 42.47 | 80.20 | 249.77 | 447.39 | 626.58 | 2,464.99 | <0.001 | <0.001 |

#### Supplementary Table 7. ALS and control participant demographics: Plasma and urine with and without genetics

##### A. Plasma + ALS-PGS

| **Covariate** | **ALS**, N = 266^1^ | **Control**, N = 179^1^ | **p-value**^2^ |
| --- | --- | --- | --- |
| Age at Sample Collection | 66 (58-73) | 62 (54-67) | **<0.001** |
| Sex |  |  | **<0.001** |
| Female | 115 (43%) | 106 (59%) |  |
| Male | 151 (57%) | 73 (41%) |  |
| Ethnicity |  |  | **0.023** |
| Hispanic or Latino | 0 (0%) | 4 (2.4%) |  |
| Not Hispanic or Latino | 263 (100%) | 164 (98%) |  |
| Missing | 3 | 11 |  |
| Race |  |  | >0.9 |
| American Indian and Alaska native | 1 (0.4%) | 0 (0%) |  |
| White or Caucasian | 264 (100%) | 179 (100%) |  |
| Missing | 1 | 0 |  |
| Military Service |  |  | 0.080 |
| Enlisted | 31 (12%) | 10 (6.6%) |  |
| Neither | 229 (88%) | 142 (93%) |  |
| Missing | 6 | 27 |  |
| Education |  |  | **<0.001** |
| High school or less | 82 (32%) | 10 (6.6%) |  |
| Bachelor’s degree | 57 (22%) | 55 (36%) |  |
| Some postsecondary | 79 (30%) | 35 (23%) |  |
| Graduate degree | 42 (16%) | 52 (34%) |  |
| Missing | 6 | 27 |  |
| Family History of ALS | 28 (11%) | 0 (0%) | **<0.001** |
| Missing | 8 | 67 |  |
| ALS PGS | 0.13 (-0.55-0.83) | 0.01 (-0.64-0.60) | 0.085 |
| Age at Diagnosis | 65 (57-72) |  |  |
| Diagnosis |  |  |  |
| ALS | 246 (92%) |  |  |
| ALS/FTD | 13 (4.9%) |  |  |
| BAD | 3 (1.1%) |  |  |
| Flail | 1 (0.4%) |  |  |
| PMA | 3 (1.1%) |  |  |
| El Escorial Criteria |  |  |  |
| Definite | 74 (29%) |  |  |
| Probable | 81 (32%) |  |  |
| Probable, lab supported | 65 (26%) |  |  |
| Possible | 33 (13%) |  |  |
| suspected | 0 (0%) |  |  |
| Missing | 13 |  |  |
| Onset Segment |  |  |  |
| Bulbar | 69 (26%) |  |  |
| Cervical | 84 (32%) |  |  |
| Lumbar | 108 (41%) |  |  |
| Respiratory | 1 (0.4%) |  |  |
| Thoracic | 3 (1.1%) |  |  |
| Missing | 1 |  |  |
| ALSFRS-R | 38 (33-41) |  |  |
| Missing | 2 |  |  |
| Time Between Symptom Onset and Diagnosis | 1.05 (0.62-1.72) |  |  |
| Missing | 1 |  |  |
| Time Between Diagnosis and Sample | 0.59 (0.30-1.06) |  |  |
| ^1^Median (25%-75%); n (%) | | | |
| ^2^Wilcoxon rank sum test; Pearson’s Chi-squared test; Fisher’s exact test | | | |
| Abbreviations: ALS, amyotrophic lateral sclerosis; ALSFRS-R, Revised ALS Functional Rating Scale; ALS-PGS, amyotrophic lateral sclerosis polygenic risk score. | | | |

##### B. Plasma + Metal-PGS

| **Covariate** | **ALS**, N = 297^1^ | **Control**, N = 179^1^ | **p-value**^2^ |
| --- | --- | --- | --- |
| Age at Sample Collection | 65 (57-73) | 62 (54-67) | **<0.001** |
| Sex |  |  | **<0.001** |
| Female | 129 (43%) | 106 (59%) |  |
| Male | 168 (57%) | 73 (41%) |  |
| Ethnicity |  |  | **0.017** |
| Hispanic or Latino | 0 (0%) | 4 (2.4%) |  |
| Not Hispanic or Latino | 292 (100%) | 164 (98%) |  |
| Missing | 5 | 11 |  |
| Race |  |  | >0.9 |
| American Indian and Alaska native | 1 (0.3%) | 0 (0%) |  |
| White or Caucasian | 295 (100%) | 179 (100%) |  |
| Missing | 1 | 0 |  |
| Military Service |  |  | 0.058 |
| Enlisted | 36 (12%) | 10 (6.6%) |  |
| Neither | 255 (88%) | 142 (93%) |  |
| Missing | 6 | 27 |  |
| Education |  |  | **<0.001** |
| High school or less | 91 (31%) | 10 (6.6%) |  |
| Bachelor’s degree | 65 (22%) | 55 (36%) |  |
| Some postsecondary | 89 (31%) | 35 (23%) |  |
| Graduate degree | 46 (16%) | 52 (34%) |  |
| Missing | 6 | 27 |  |
| Family History of ALS | 31 (11%) | 0 (0%) | **<0.001** |
| Missing | 9 | 67 |  |
| Metal PGS | 0.37 (0.34-0.40) | 0.37 (0.35-0.40) | 0.8 |
| Age at Diagnosis | 64 (56-72) |  |  |
| Diagnosis |  |  |  |
| ALS | 276 (93%) |  |  |
| ALS/FTD | 13 (4.4%) |  |  |
| BAD | 4 (1.3%) |  |  |
| Flail | 1 (0.3%) |  |  |
| PMA | 3 (1.0%) |  |  |
| El Escorial Criteria |  |  |  |
| Definite | 81 (29%) |  |  |
| Probable | 89 (31%) |  |  |
| Probable, lab supported | 76 (27%) |  |  |
| Possible | 37 (13%) |  |  |
| suspected | 0 (0%) |  |  |
| Missing | 14 |  |  |
| Onset Segment |  |  |  |
| Bulbar | 75 (25%) |  |  |
| Cervical | 101 (34%) |  |  |
| Lumbar | 116 (39%) |  |  |
| Respiratory | 1 (0.3%) |  |  |
| Thoracic | 3 (1.0%) |  |  |
| Missing | 1 |  |  |
| ALSFRS-R | 38 (33-41) |  |  |
| Missing | 2 |  |  |
| Time Between Symptom Onset and Diagnosis | 1.07 (0.66-1.84) |  |  |
| Missing | 1 |  |  |
| Time Between Diagnosis and Sample | 0.65 (0.32-1.27) |  |  |
| ^1^Median (25%-75%); n (%) | | | |
| ^2^Wilcoxon rank sum test; Pearson’s Chi-squared test; Fisher’s exact test | | | |
| Abbreviations: ALS, amyotrophic lateral sclerosis; ALSFRS-R, Revised ALS Functional Rating Scale; Metal-PGS, metal polygenic risk score. | | | |

##### C. Urine + ALS-PGS

| **Covariate** | **ALS**, N = 243^1^ | **Control**, N = 135^1^ | **p-value**^2^ |
| --- | --- | --- | --- |
| Age at Sample Collection | 66 (58-72) | 62 (55-67) | **<0.001** |
| Sex |  |  | **<0.001** |
| Female | 94 (39%) | 81 (60%) |  |
| Male | 149 (61%) | 54 (40%) |  |
| Ethnicity |  |  | **0.042** |
| Hispanic or Latino | 0 (0%) | 3 (2.3%) |  |
| Not Hispanic or Latino | 241 (100%) | 126 (98%) |  |
| Missing | 2 | 6 |  |
| Race |  |  | >0.9 |
| American Indian and Alaska native | 1 (0.4%) | 0 (0%) |  |
| White or Caucasian | 241 (100%) | 135 (100%) |  |
| Missing | 1 | 0 |  |
| Military Service |  |  | 0.2 |
| Enlisted | 28 (12%) | 8 (7.0%) |  |
| Neither | 211 (88%) | 107 (93%) |  |
| Missing | 4 | 20 |  |
| Education |  |  | **<0.001** |
| High school or less | 64 (27%) | 6 (5.2%) |  |
| Bachelor’s degree | 56 (23%) | 42 (37%) |  |
| Some postsecondary | 77 (32%) | 22 (19%) |  |
| Graduate degree | 42 (18%) | 45 (39%) |  |
| Missing | 4 | 20 |  |
| Family History of ALS | 23 (9.7%) | 0 (0%) | **0.002** |
| Missing | 5 | 39 |  |
| ALS PGS | 0.17 (-0.44-0.88) | 0.01 (-0.65-0.57) | **0.009** |
| Age at Diagnosis | 65 (57-72) |  |  |
| Diagnosis |  |  |  |
| ALS | 220 (91%) |  |  |
| ALS/FTD | 12 (4.9%) |  |  |
| BAD | 8 (3.3%) |  |  |
| PMA | 3 (1.2%) |  |  |
| El Escorial Criteria |  |  |  |
| Definite | 60 (27%) |  |  |
| Probable | 72 (32%) |  |  |
| Probable, lab supported | 62 (27%) |  |  |
| Possible | 32 (14%) |  |  |
| suspected | 0 (0%) |  |  |
| Missing | 17 |  |  |
| Onset segment |  |  |  |
| Bulbar | 66 (27%) |  |  |
| Cervical | 86 (36%) |  |  |
| Lumbar | 87 (36%) |  |  |
| Respiratory | 1 (0.4%) |  |  |
| Thoracic | 2 (0.8%) |  |  |
| Missing | 1 |  |  |
| ALSFRS-R | 38.0 (34.0-41.0) |  |  |
| Missing | 1 |  |  |
| Time Between Symptom Onset and Diagnosis | 1.06 (0.67-1.72) |  |  |
| Missing | 1 |  |  |
| Time Between Diagnosis and Sample | 0.52 (0.30-0.86) |  |  |
| ^1^Median (25%-75%); n (%) | | | |
| ^2^Wilcoxon rank sum test; Pearson’s Chi-squared test; Fisher’s exact test | | | |
| Abbreviations: ALS, amyotrophic lateral sclerosis; ALSFRS-R, Revised ALS Functional Rating Scale; ALS-PGS, amyotrophic lateral sclerosis polygenic risk score. | | | |

##### D. Urine + Metal-PGS

| **Covariate** | **ALS**, N = 248^1^ | **Control**, N = 135^1^ | **p-value**^2^ |
| --- | --- | --- | --- |
| Age at Sample Collection | 65 (57-72) | 62 (55-67) | **<0.001** |
| Sex |  |  | **<0.001** |
| Female | 94 (38%) | 81 (60%) |  |
| Male | 154 (62%) | 54 (40%) |  |
| Ethnicity |  |  | **0.040** |
| Hispanic or Latino | 0 (0%) | 3 (2.3%) |  |
| Not Hispanic or Latino | 246 (100%) | 126 (98%) |  |
| Missing | 2 | 6 |  |
| Race |  |  | >0.9 |
| American Indian and Alaska native | 1 (0.4%) | 0 (0%) |  |
| White or Caucasian | 246 (100%) | 135 (100%) |  |
| Missing | 1 | 0 |  |
| Military Service |  |  | 0.15 |
| Enlisted | 29 (12%) | 8 (7.0%) |  |
| Neither | 213 (88%) | 107 (93%) |  |
| Missing | 6 | 20 |  |
| Education |  |  | **<0.001** |
| High school or less | 65 (27%) | 6 (5.2%) |  |
| Bachelor’s degree | 58 (24%) | 42 (37%) |  |
| Some postsecondary | 77 (32%) | 22 (19%) |  |
| Graduate degree | 42 (17%) | 45 (39%) |  |
| Missing | 6 | 20 |  |
| Family History of ALS | 23 (9.5%) | 0 (0%) | **0.002** |
| Missing | 5 | 39 |  |
| Metal PGS | 0.37 (0.35-0.40) | 0.37 (0.35-0.40) | 0.5 |
| Age at Diagnosis | 65 (56-72) |  |  |
| Diagnosis |  |  |  |
| ALS | 225 (91%) |  |  |
| ALS/FTD | 12 (4.8%) |  |  |
| BAD | 8 (3.2%) |  |  |
| PMA | 3 (1.2%) |  |  |
| El Escorial Criteria |  |  |  |
| Definite | 62 (27%) |  |  |
| Probable | 73 (32%) |  |  |
| Probable, lab supported | 64 (28%) |  |  |
| Possible | 32 (14%) |  |  |
| suspected | 0 (0%) |  |  |
| Missing | 17 |  |  |
| Onset Segment |  |  |  |
| Bulbar | 66 (27%) |  |  |
| Cervical | 89 (36%) |  |  |
| Lumbar | 89 (36%) |  |  |
| Respiratory | 1 (0.4%) |  |  |
| Thoracic | 2 (0.8%) |  |  |
| Missing | 1 |  |  |
| ALSFRS-R | 38.0 (34.0-41.0) |  |  |
| Missing | 1 |  |  |
| Time Between Symptom Onset and Diagnosis | 1.06 (0.67-1.74) |  |  |
| Missing | 1 |  |  |
| Time Between Diagnosis and Sample | 0.53 (0.31-0.89) |  |  |
| ^1^Median (25%-75%); n (%) | | | |
| ^2^Wilcoxon rank sum test; Pearson’s Chi-squared test; Fisher’s exact test | | | |
| Abbreviations: ALS, amyotrophic lateral sclerosis; ALSFRS-R, Revised ALS Functional Rating Scale; Metal-PGS, metal polygenic risk score. | | | |

#### Supplementary Table 8. Supplement and/or vitamin usage: Plasma

Abbreviations: N, the number of subjects in the group; SD, standard deviation; Q10, 10^th^ Percentile; Q25, 25^th^ Percentile; Q50, median; Q75, 75^th^ Percentile; Q90, 90^th^ Percentile. P-value, the *p*-value from the Wilcoxon rank sum test compares metal levels between the case and control groups; BH, Benjamini-Hochberg; %BDL, the percentage of samples below the detection limit. Red, median value is higher in supplement and/or vitamin use group; significance based on *q*-value.

|  | | **Supplement / Vitamin Use Positive, N=349  (μg/ℓ)** | | | | | | | | | **Supplement / Vitamin Use Negative, N=319  (μg/ℓ)** | | | | | | | | |  | |
| --- | --- | --- | --- | --- | --- | --- | --- | --- | --- | --- | --- | --- | --- | --- | --- | --- | --- | --- | --- | --- | --- |
| **Metal** | **%BDL** | **Mean** | **SD** | **Min** | **Q10** | **Q25** | **Q50** | **Q75** | **Q90** | **Max** | **Mean** | **SD** | **Min** | **Q10** | **Q25** | **Q50** | **Q75** | **Q90** | **Max** | **P-value** | **Q-value (BH)** |
| Aluminum | 84.0% | 42.18 | 284.91 | 14.14 | 14.14 | 14.14 | 14.14 | 14.14 | 22.37 | 4,612.83 | 70.73 | 582.33 | 14.14 | 14.14 | 14.14 | 14.14 | 14.14 | 25.63 | 9,595.32 | 0.673 | >0.999 |
| Antimony | 3.5% | 3.26 | 2.19 | 0.08 | 1.91 | 2.45 | 3.15 | 3.77 | 4.56 | 28.02 | 3.50 | 1.77 | 0.08 | 2.22 | 2.63 | 3.20 | 3.96 | 5.20 | 23.36 | 0.032 | 0.574 |
| Arsenic | 0.1% | 0.62 | 1.75 | 0.09 | 0.15 | 0.20 | 0.27 | 0.43 | 1.19 | 29.29 | 0.58 | 0.93 | 0.09 | 0.17 | 0.20 | 0.28 | 0.53 | 1.20 | 8.48 | 0.142 | >0.999 |
| Barium | 0.1% | 1.72 | 1.53 | 0.22 | 0.57 | 0.81 | 1.37 | 2.07 | 3.13 | 15.42 | 2.26 | 5.69 | 0.22 | 0.54 | 0.91 | 1.53 | 2.30 | 3.44 | 95.58 | 0.049 | 0.826 |
| Beryllium | 99.6% | 0.07 | 0.00 | 0.07 | 0.07 | 0.07 | 0.07 | 0.07 | 0.07 | 0.11 | 0.07 | 0.02 | 0.07 | 0.07 | 0.07 | 0.07 | 0.07 | 0.07 | 0.34 | 0.512 | >0.999 |
| Cadmium | 36.3% | 0.01 | 0.01 | 0.00 | 0.00 | 0.00 | 0.01 | 0.02 | 0.03 | 0.09 | 0.01 | 0.02 | 0.00 | 0.00 | 0.00 | 0.01 | 0.02 | 0.03 | 0.25 | 0.400 | >0.999 |
| Chromium | 0.4% | 0.67 | 2.25 | 0.06 | 0.23 | 0.29 | 0.37 | 0.51 | 0.83 | 39.08 | 0.52 | 0.97 | 0.09 | 0.24 | 0.29 | 0.35 | 0.46 | 0.58 | 11.65 | 0.279 | >0.999 |
| Cobalt | 0.1% | 0.38 | 1.09 | 0.11 | 0.14 | 0.16 | 0.20 | 0.27 | 0.40 | 13.40 | 0.28 | 0.35 | 0.09 | 0.14 | 0.17 | 0.19 | 0.26 | 0.40 | 3.89 | 0.818 | >0.999 |
| Copper | 0.1% | 1,096.36 | 222.91 | 587.71 | 848.56 | 948.78 | 1,065.06 | 1,209.92 | 1,353.82 | 2,432.46 | 1,089.22 | 207.60 | 610.65 | 854.84 | 940.34 | 1,059.09 | 1,205.86 | 1,359.38 | 1,894.29 | 0.731 | >0.999 |
| Iron | 0.1% | 1,322.66 | 831.47 | 237.89 | 732.10 | 912.50 | 1,189.09 | 1,434.84 | 1,935.70 | 9,084.29 | 1,315.77 | 644.33 | 386.48 | 775.08 | 938.31 | 1,189.46 | 1,554.31 | 1,916.29 | 7,109.31 | 0.341 | >0.999 |
| Lead | 0.8% | 0.43 | 0.63 | 0.03 | 0.08 | 0.14 | 0.24 | 0.43 | 0.91 | 6.36 | 0.42 | 1.04 | 0.02 | 0.08 | 0.13 | 0.21 | 0.35 | 0.72 | 16.40 | 0.025 | 0.473 |
| Manganese | 0.1% | 1.51 | 4.33 | 0.24 | 0.40 | 0.47 | 0.55 | 0.71 | 4.24 | 70.17 | 1.16 | 2.15 | 0.29 | 0.40 | 0.49 | 0.59 | 0.75 | 1.57 | 21.91 | 0.134 | >0.999 |
| Mercury | 85.2% | 1.16 | 0.90 | 0.88 | 0.88 | 0.88 | 0.88 | 0.88 | 1.65 | 8.02 | 1.29 | 1.62 | 0.88 | 0.88 | 0.88 | 0.88 | 0.88 | 1.84 | 16.50 | 0.666 | >0.999 |
| Molybdenum | 85.0% | 2.12 | 1.02 | 1.75 | 1.75 | 1.75 | 1.75 | 1.75 | 2.98 | 7.66 | 2.06 | 1.03 | 1.75 | 1.75 | 1.75 | 1.75 | 1.75 | 2.98 | 12.01 | 0.279 | >0.999 |
| Nickel | 7.7% | 0.80 | 0.55 | 0.20 | 0.32 | 0.51 | 0.77 | 0.96 | 1.21 | 8.56 | 0.97 | 1.21 | 0.20 | 0.38 | 0.57 | 0.85 | 1.06 | 1.30 | 17.28 | 0.006 | 0.119 |
| Selenium | 0.1% | 141.47 | 29.00 | 70.80 | 117.92 | 125.96 | 137.90 | 149.58 | 168.09 | 361.30 | 135.09 | 19.44 | 86.32 | 114.18 | 123.23 | 134.63 | 143.74 | 153.92 | 266.83 | 0.002 | 0.041 |
| Silver | 17.9% | 0.14 | 0.33 | 0.02 | 0.02 | 0.04 | 0.07 | 0.13 | 0.27 | 5.37 | 0.13 | 0.17 | 0.02 | 0.02 | 0.04 | 0.08 | 0.16 | 0.27 | 2.15 | 0.136 | >0.999 |
| Strontium | 0.1% | 26.99 | 18.87 | 9.07 | 15.26 | 18.82 | 23.63 | 29.62 | 39.29 | 254.24 | 27.42 | 12.65 | 8.14 | 15.71 | 19.38 | 25.04 | 31.96 | 40.55 | 152.55 | 0.067 | >0.999 |
| Thallium | 2.8% | 0.03 | 0.01 | 0.01 | 0.01 | 0.02 | 0.03 | 0.04 | 0.05 | 0.11 | 0.03 | 0.01 | 0.01 | 0.01 | 0.02 | 0.03 | 0.04 | 0.05 | 0.09 | 0.211 | >0.999 |
| Tin | 19.3% | 0.35 | 0.89 | 0.07 | 0.07 | 0.12 | 0.20 | 0.34 | 0.58 | 15.05 | 0.32 | 0.60 | 0.07 | 0.07 | 0.11 | 0.18 | 0.32 | 0.55 | 6.88 | 0.383 | >0.999 |
| Uranium | 98.6% | 0.02 | 0.01 | 0.02 | 0.02 | 0.02 | 0.02 | 0.02 | 0.02 | 0.18 | 0.02 | 0.02 | 0.02 | 0.02 | 0.02 | 0.02 | 0.02 | 0.02 | 0.42 | 0.160 | >0.999 |
| Vanadium | 0.3% | 0.11 | 0.49 | 0.02 | 0.03 | 0.04 | 0.05 | 0.07 | 0.12 | 8.14 | 0.11 | 0.53 | 0.01 | 0.03 | 0.04 | 0.04 | 0.06 | 0.10 | 8.36 | <0.001 | 0.017 |
| Zinc | 0.1% | 819.19 | 191.05 | 429.59 | 636.19 | 696.36 | 776.72 | 902.62 | 1,052.09 | 1,724.57 | 790.23 | 194.98 | 456.70 | 612.69 | 676.35 | 756.36 | 857.12 | 960.27 | 2,217.53 | 0.017 | 0.344 |

#### Supplementary Table 9. Supplement and/or vitamin usage: Urine

Abbreviations: N, the number of subjects in the group; SD, standard deviation; Q10, 10^th^ Percentile; Q25, 25^th^ Percentile; Q50, median; Q75, 75^th^ Percentile; Q90, 90^th^ Percentile. P-value, the p-value from the Wilcoxon rank sum test compares metal levels between the case and control groups.; BH, Benjamini-Hochberg; %BDL, the percentage of samples below the detection limit.

|  | | **Supplement / Vitamin Use Positive, N=294  (μg/ℓ)** | | | | | | | | | **Supplement / Vitamin Use Negative, N=222  (μg/ℓ)** | | | | | | | | |  | |
| --- | --- | --- | --- | --- | --- | --- | --- | --- | --- | --- | --- | --- | --- | --- | --- | --- | --- | --- | --- | --- | --- |
| **Metal** | **%BDL** | **Mean** | **SD** | **Min** | **Q10** | **Q25** | **Q50** | **Q75** | **Q90** | **Max** | **Mean** | **SD** | **Min** | **Q10** | **Q25** | **Q50** | **Q75** | **Q90** | **Max** | **P-value** | **Q-value (BH)** |
| Aluminum | 35.0% | 13.89 | 35.09 | 2.84 | 2.84 | 2.84 | 5.68 | 10.10 | 20.88 | 364.19 | 13.99 | 36.25 | 2.84 | 2.84 | 2.84 | 5.13 | 10.12 | 20.31 | 279.19 | 0.397 | >0.999 |
| Antimony | 37.8% | 0.07 | 0.51 | 0.01 | 0.01 | 0.01 | 0.02 | 0.05 | 0.10 | 8.73 | 0.04 | 0.05 | 0.01 | 0.01 | 0.01 | 0.02 | 0.06 | 0.10 | 0.45 | 0.664 | >0.999 |
| Arsenic | 0.2% | 23.14 | 131.11 | 0.22 | 2.28 | 3.81 | 6.99 | 14.60 | 32.72 | 2,212.14 | 15.01 | 25.72 | 1.19 | 2.12 | 3.79 | 7.51 | 14.56 | 32.47 | 239.70 | 0.878 | >0.999 |
| Barium | 0.0% | 3.34 | 14.28 | 0.11 | 0.41 | 0.81 | 1.73 | 2.97 | 5.91 | 242.05 | 2.32 | 3.21 | 0.04 | 0.31 | 0.64 | 1.41 | 2.67 | 4.99 | 32.75 | 0.085 | >0.999 |
| Beryllium | 92.3% | 0.02 | 0.00 | 0.02 | 0.02 | 0.02 | 0.02 | 0.02 | 0.02 | 0.05 | 0.02 | 0.01 | 0.02 | 0.02 | 0.02 | 0.02 | 0.02 | 0.03 | 0.07 | 0.039 | 0.679 |
| Cadmium | 0.0% | 0.43 | 0.46 | 0.02 | 0.08 | 0.16 | 0.28 | 0.51 | 0.90 | 3.45 | 0.41 | 0.47 | 0.01 | 0.08 | 0.14 | 0.27 | 0.52 | 0.85 | 3.77 | 0.427 | >0.999 |
| Chromium | 79.9% | 0.88 | 1.31 | 0.53 | 0.53 | 0.53 | 0.53 | 0.53 | 1.19 | 16.13 | 0.86 | 1.79 | 0.53 | 0.53 | 0.53 | 0.53 | 0.53 | 1.03 | 18.40 | 0.377 | >0.999 |
| Cobalt | 0.2% | 0.65 | 2.63 | 0.01 | 0.07 | 0.11 | 0.18 | 0.30 | 0.66 | 36.65 | 0.43 | 1.33 | 0.01 | 0.06 | 0.10 | 0.20 | 0.35 | 0.77 | 18.74 | 0.340 | >0.999 |
| Copper | 0.2% | 10.41 | 7.27 | 0.33 | 3.06 | 5.07 | 8.69 | 14.02 | 18.69 | 57.21 | 13.31 | 27.47 | 0.68 | 2.17 | 4.32 | 8.30 | 13.02 | 21.39 | 277.36 | 0.263 | >0.999 |
| Iron | 26.3% | 12.26 | 11.61 | 3.07 | 3.07 | 4.77 | 8.70 | 15.81 | 24.88 | 78.12 | 16.68 | 38.87 | 3.07 | 3.07 | 3.07 | 7.24 | 13.12 | 28.84 | 382.91 | 0.025 | 0.510 |
| Lead | 14.5% | 0.19 | 0.12 | 0.04 | 0.04 | 0.10 | 0.16 | 0.24 | 0.35 | 0.85 | 0.20 | 0.15 | 0.04 | 0.04 | 0.09 | 0.18 | 0.26 | 0.35 | 1.08 | 0.443 | >0.999 |
| Manganese | 49.9% | 0.21 | 0.44 | 0.06 | 0.06 | 0.06 | 0.09 | 0.16 | 0.39 | 3.89 | 0.16 | 0.30 | 0.06 | 0.06 | 0.06 | 0.06 | 0.14 | 0.29 | 3.15 | 0.010 | 0.237 |
| Mercury | 77.0% | 0.39 | 0.27 | 0.28 | 0.28 | 0.28 | 0.28 | 0.28 | 0.67 | 2.76 | 0.40 | 0.26 | 0.28 | 0.28 | 0.28 | 0.28 | 0.28 | 0.71 | 1.93 | 0.472 | >0.999 |
| Molybdenum | 0.0% | 58.67 | 57.10 | 2.47 | 10.64 | 22.86 | 39.68 | 72.86 | 126.73 | 423.66 | 49.69 | 49.41 | 1.96 | 9.43 | 16.51 | 34.91 | 63.13 | 105.81 | 331.18 | 0.032 | 0.608 |
| Nickel | 17.7% | 1.51 | 1.61 | 0.18 | 0.18 | 0.47 | 1.13 | 1.95 | 3.23 | 12.23 | 1.74 | 2.28 | 0.18 | 0.18 | 0.38 | 1.05 | 1.97 | 4.14 | 15.04 | 0.836 | >0.999 |
| Selenium | 0.0% | 51.87 | 47.39 | 2.95 | 11.06 | 22.25 | 37.28 | 64.04 | 100.67 | 353.37 | 41.57 | 31.38 | 2.52 | 10.14 | 17.50 | 33.28 | 55.35 | 79.25 | 199.74 | 0.038 | 0.679 |
| Silver | 98.0% | 0.02 | 0.09 | 0.02 | 0.02 | 0.02 | 0.02 | 0.02 | 0.02 | 1.51 | 0.02 | 0.00 | 0.02 | 0.02 | 0.02 | 0.02 | 0.02 | 0.02 | 0.05 | 0.045 | 0.722 |
| Strontium | 0.0% | 147.16 | 142.81 | 10.74 | 36.68 | 67.63 | 111.91 | 174.59 | 277.71 | 1,351.11 | 131.12 | 98.38 | 5.48 | 32.32 | 57.36 | 114.16 | 180.96 | 240.38 | 700.56 | 0.540 | >0.999 |
| Thallium | 32.2% | 0.25 | 0.33 | 0.04 | 0.04 | 0.04 | 0.14 | 0.31 | 0.64 | 2.65 | 0.28 | 0.32 | 0.04 | 0.04 | 0.04 | 0.17 | 0.38 | 0.70 | 1.98 | 0.301 | >0.999 |
| Tin | 6.4% | 1.55 | 9.17 | 0.02 | 0.07 | 0.16 | 0.38 | 0.91 | 1.78 | 149.69 | 1.10 | 2.95 | 0.02 | 0.06 | 0.16 | 0.35 | 0.89 | 2.03 | 29.13 | 0.858 | >0.999 |
| Uranium | 0.0% | 1.25 | 9.27 | 0.03 | 0.14 | 0.24 | 0.40 | 0.78 | 1.08 | 152.48 | 0.57 | 0.57 | 0.02 | 0.10 | 0.21 | 0.40 | 0.70 | 1.20 | 4.13 | 0.490 | >0.999 |
| Vanadium | 35.2% | 0.14 | 0.98 | 0.02 | 0.02 | 0.02 | 0.05 | 0.10 | 0.18 | 16.88 | 0.10 | 0.25 | 0.02 | 0.02 | 0.02 | 0.04 | 0.09 | 0.16 | 2.97 | 0.013 | 0.282 |
| Zinc | 0.2% | 538.48 | 523.61 | 5.94 | 82.48 | 197.11 | 391.16 | 718.75 | 1,082.71 | 4,502.51 | 520.85 | 787.13 | 12.12 | 54.15 | 148.79 | 345.95 | 582.89 | 956.46 | 6,717.82 | 0.020 | 0.420 |

### SUPPLEMENTAL FIGURES

#### Supplemental Figure 1. Single nucleotide polymorphism (SNP) filtering based on quality control assessments and post imputation SNP filtering.

HWE, Hardy-Weinberg equilibrium; MAF, minor allele frequency.

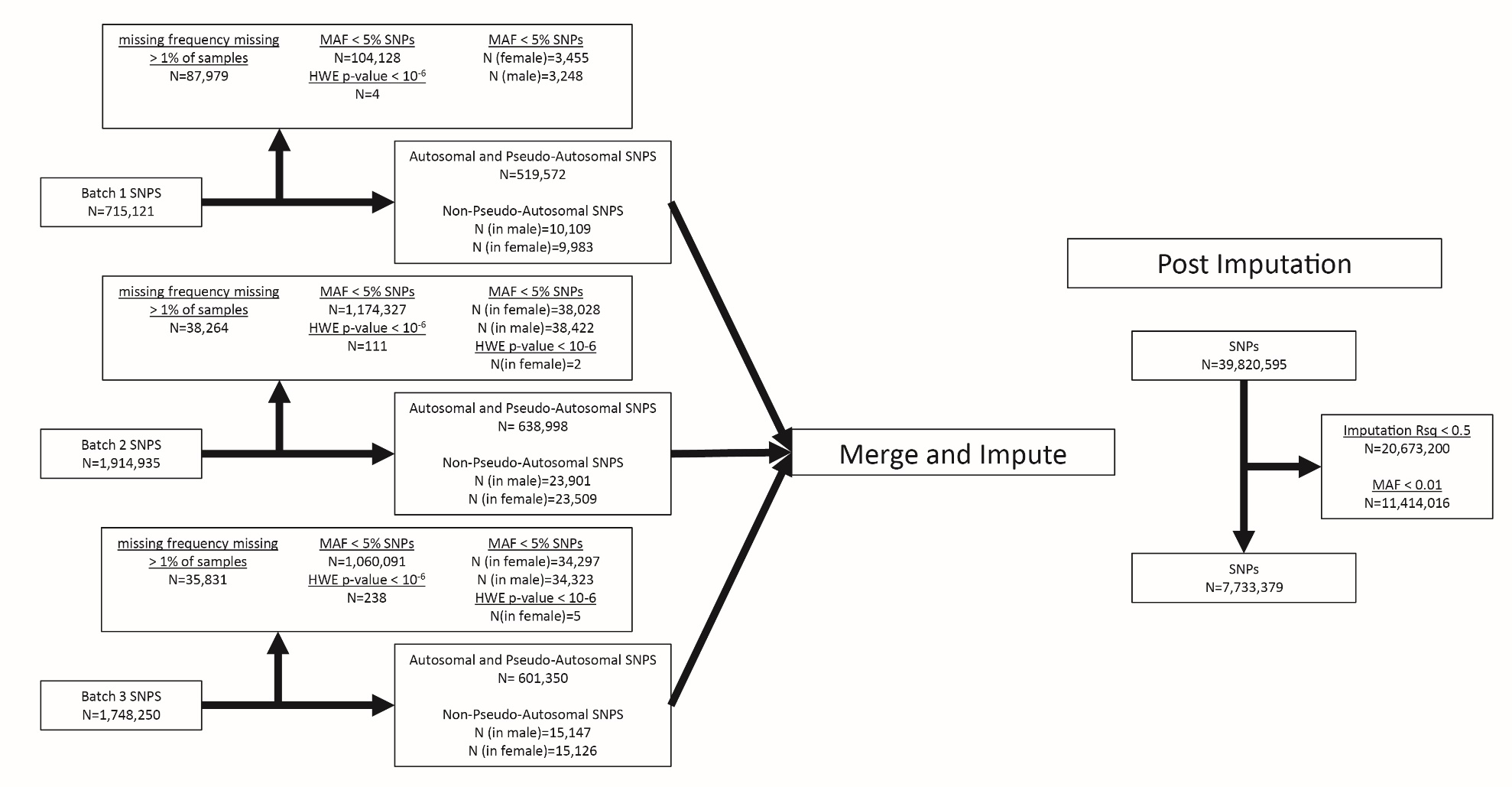

#### Supplemental Figure 2. Genetic sample filtering based on quality control assessments.

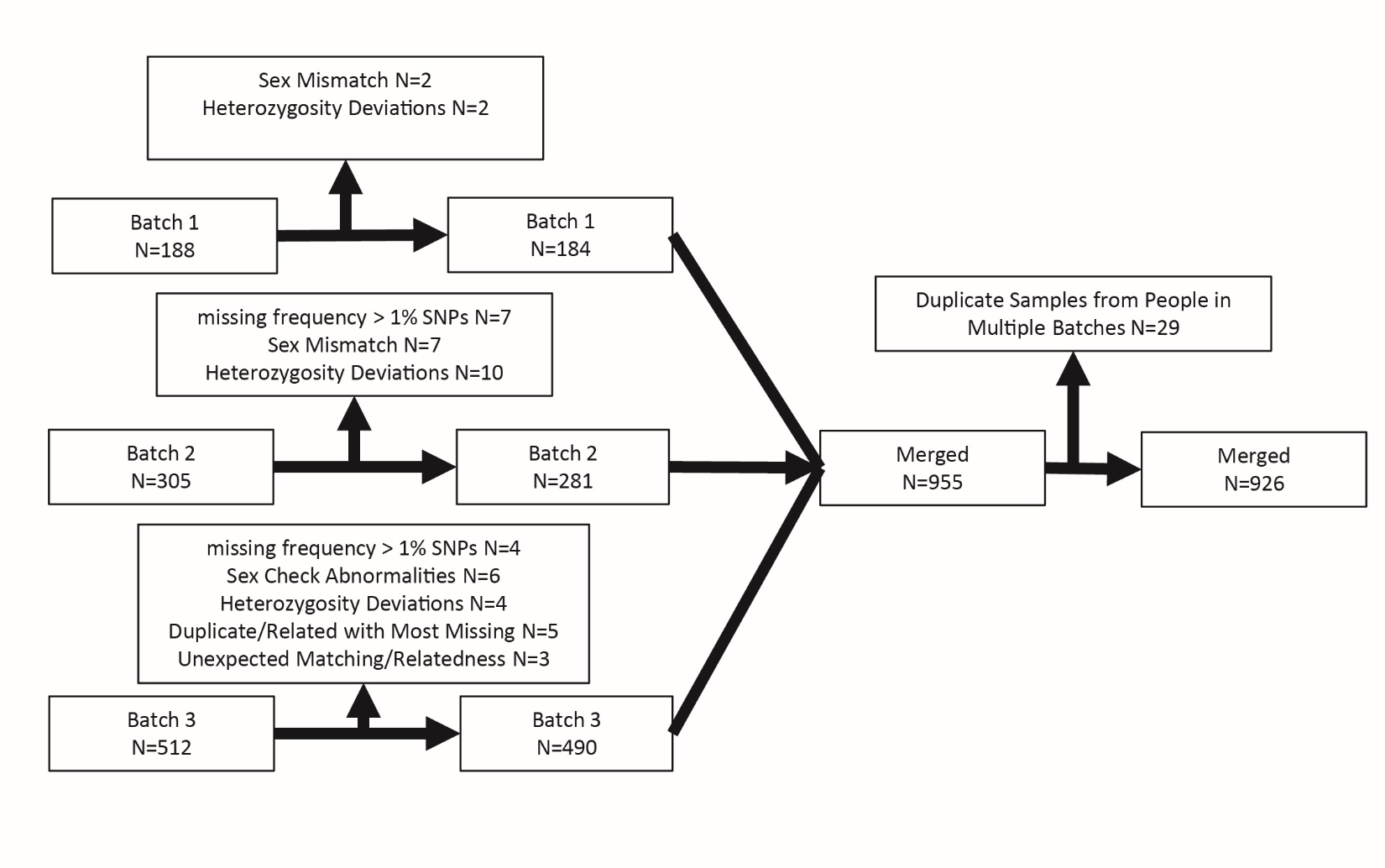

#### Supplemental Figure 3. Metal correlations for plasma and urine samples.

Spearman correlations of log-transformed metal levels in (**A**) plasma, (**B**) urine, and (**C**) between biofluids. **p*-value < 0.05; ***p*-value < 0.01; ****p*-value < 0.001; *****p*-value < 0.0001. Red, positive correlation; Blue, negative correlation.

##### A. Plasma

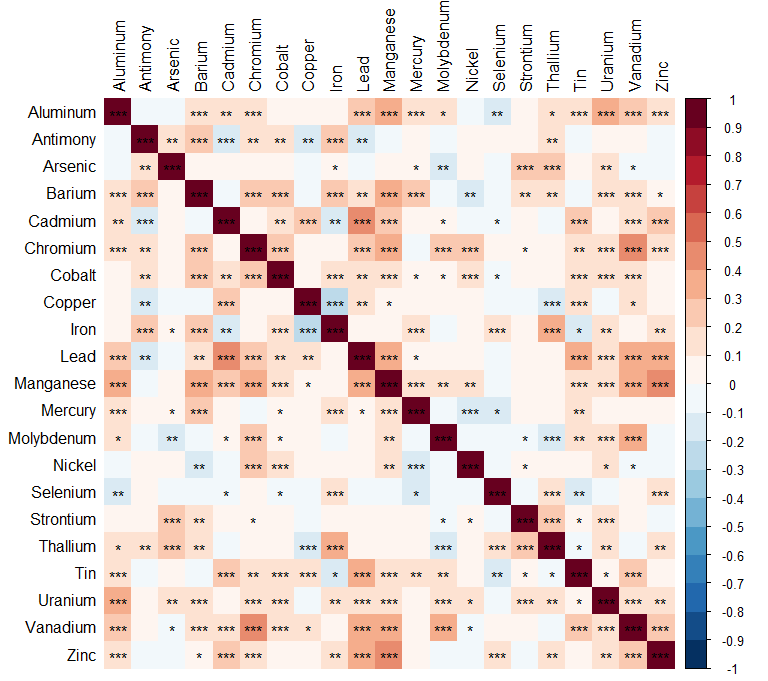

##### B. Urine

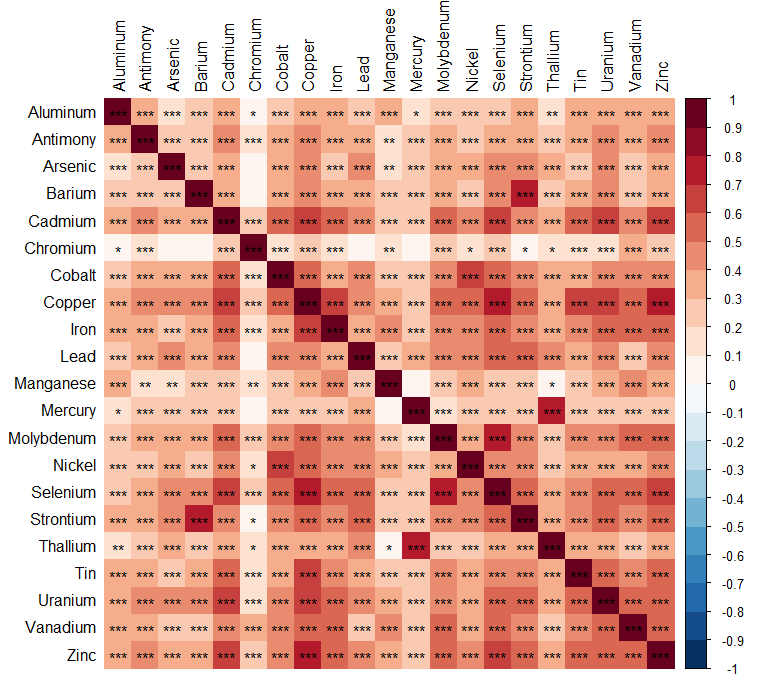

##### C. Plasma and Urine

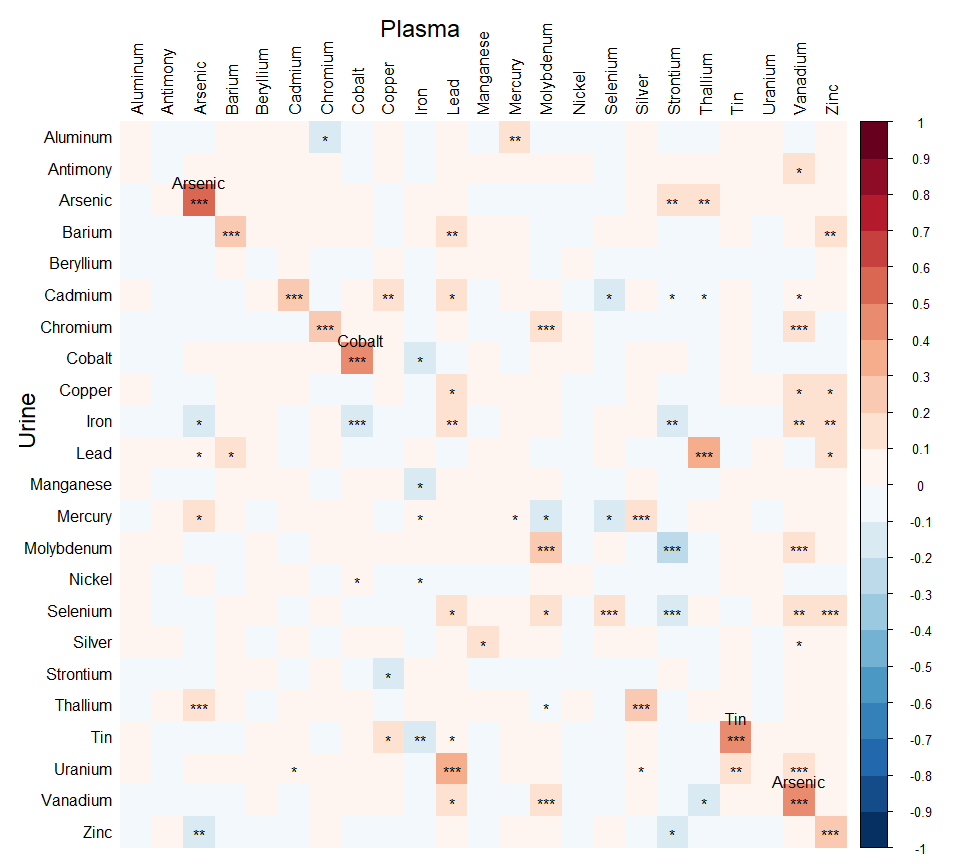

#### Supplemental Figure 4. Environmental risk score (ERS) risk heatmaps.

Heatmaps of (A) plasma and (B) urine metal levels. Rows represent standardized log-transformed metal levels, while columns represent subjects categorized by sex (F, female; M, male) and amyotrophic lateral sclerosis (ALS) cases/controls. Within each category, the arrangement is based on environmental risk scores (plasma, ERS^PR^; urine, ERS^UR^) derived from ALS case/control logistic regression analysis and standardized by the mean and standard deviation of the control distribution. Z-score, log-transformed and standardized metal levels.

##### A. Plasma

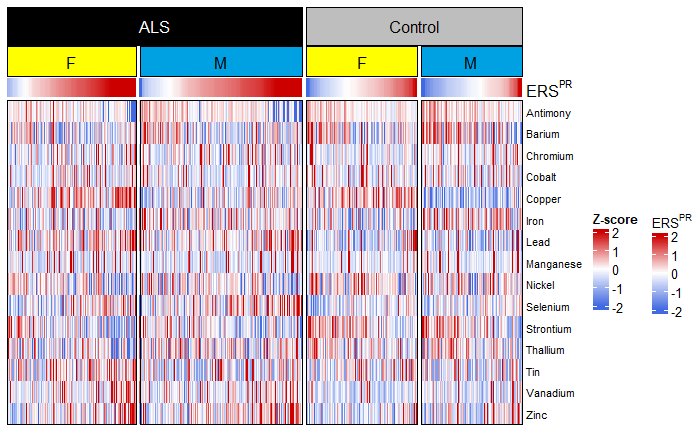

##### B. Urine

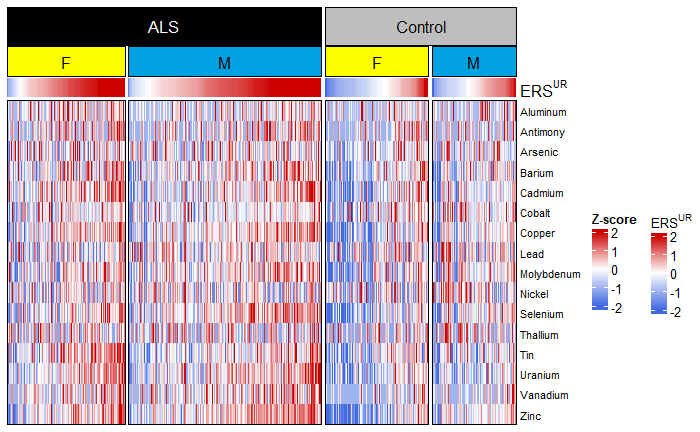

#### Supplemental Figure 5. Plasma single metal and mixture association with amyotrophic lateral sclerosis (ALS) risk and ALS polygenic risk score (ALS-PGS) forest plot.

Single metal logistic regression models based on plasma samples where the outcome is the case/control status, the predictors are metal levels, ALS-PGS, and the interaction between metal levels and ALS-PGS, and the covariates are continuous age at diagnosis, sex, military service, and the first five genetic principal components. OR 95% CI, odds ratio and its 95% confidence interval of the metal main effect; P-value, the *p*-value of metal main effect; BH, Benjamini-Hochberg; OR 95% CI (int), odds ratio and its 95% confidence interval of the interaction effect; P-value (int), the *p*-value of the interaction effect, red for significant interactions; %BDL, the percentage of samples below detection limit; ERS^PR^, plasma risk environmental risk score.

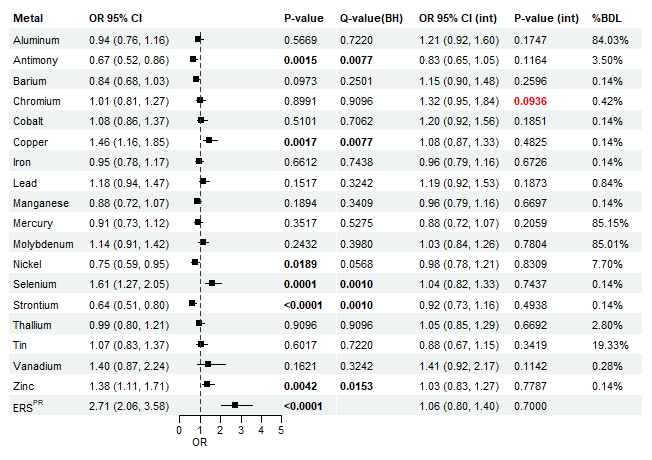

#### Supplemental Figure 6. Plasma single metal and mixture associations with amyotrophic lateral sclerosis (ALS) risk and metal polygenic risk score (metal-PGS) forest plot.

Singel metal logistic regression models based on plasma samples where the outcome is the case/control status, the predictors are metal levels, metal-PGS, and the interaction between metal levels and metal-PGS, and the covariates are continuous age at diagnosis, sex, military service, and the first five genetic principal components. OR 95% CI, odds ratio and its 95% confidence interval of the metal main effect; P-value, the *p*-value of metal main effect; BH, Benjamini-Hochberg; OR 95% CI (int), odds ratio and its 95% confidence interval of the interaction effect; P-value (int), the *p*-value of the interaction effect, red for significant interactions; %BDL, the percentage of samples below detection limit; ERS^PR^, plasma risk environmental risk score.

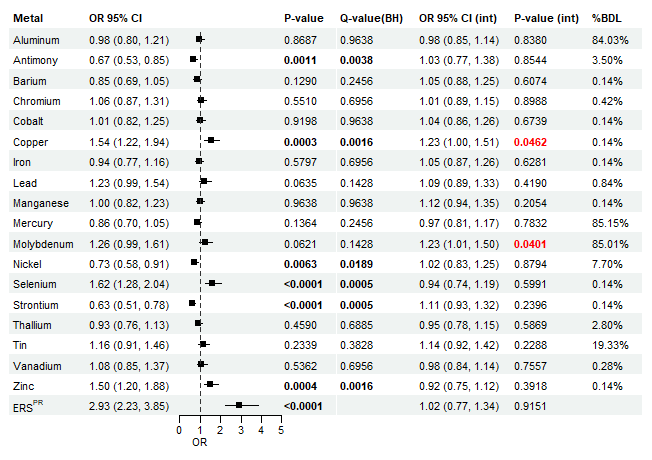

#### Supplemental Figure 7. Urine single metal and mixture associations with amyotrophic lateral sclerosis (ALS) risk and ALS polygenic risk score (ALS-PGS) forest plot.

Single metal logistic regression models based on urine samples where the outcome is the case/control status, the predictors are metal levels, ALS-PGS, and the interaction between metal levels and ALS-PGS, and the covariates are continuous age at diagnosis, sex, military service, and the first five genetic principal components. OR 95% CI, odds ratio and its 95% confidence interval of the metal main effect; P-value, the *p*-value of metal main effect; BH, Benjamini-Hochberg; OR 95% CI (int), odds ratio and its 95% confidence interval of the interaction effect; P-value (int), the *p*-value of the interaction effect, red for significant interactions; %BDL, the percentage of samples below detection limit; ERS^UR^, urine risk environmental risk score.

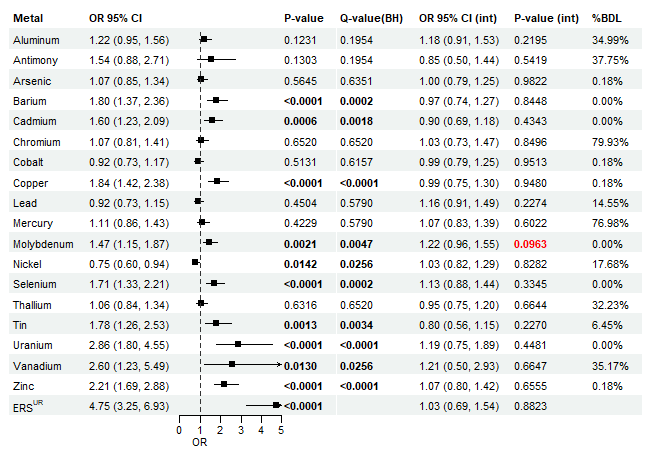

#### Supplemental Figure 8. Urine single metal and mixture associations with amyotrophic (ALS) risk and metal polygenic risk score (metal-PGS) forest plot.

Single metal logistic regression models based on urine samples where the outcome is the case/control status, the predictors are metal levels, metal-PGS, and the interaction between metal levels and metal-PGS, and the covariates are continuous age at diagnosis, sex, military service, and the first five genetic principal components. OR 95% CI, odds ratio and its 95% confidence interval of the metal main effect; P-value, the *p*-value of metal main effect; BH, Benjamini-Hochberg; OR 95% CI (int), odds ratio and its 95% confidence interval of the interaction effect; P-value (int), the *p*-value of the interaction effect, red for significant interactions; %BDL, the percentage of samples below detection limit; ERS^UR^, urine risk environmental risk score.

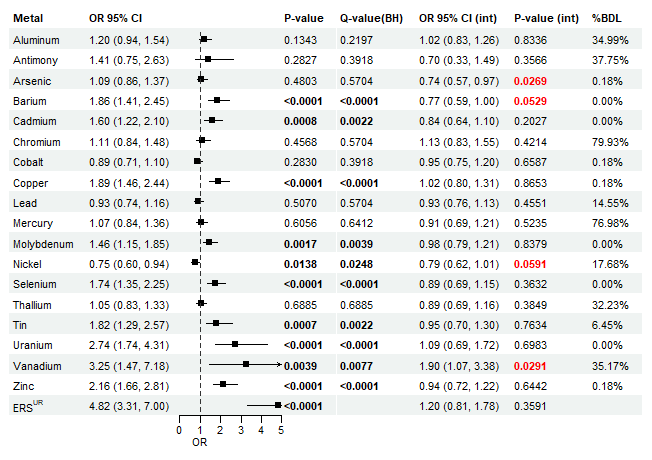

#### Supplemental Figure 9. Environmental risk score (ERS) survival heatmaps.

Heatmaps of (A) plasma and (B) urine metal levels. Rows represent standardized log-transformed metal levels, while columns represent subjects categorized by sex (F, female; M, male). Within each category, the arrangement is based on standardized environmental risk scores (plasma, ERS^PS^; urine, ERS^US^) from survival analysis. Z-score, log-transformed and standardized metal levels.

##### A. Plasma

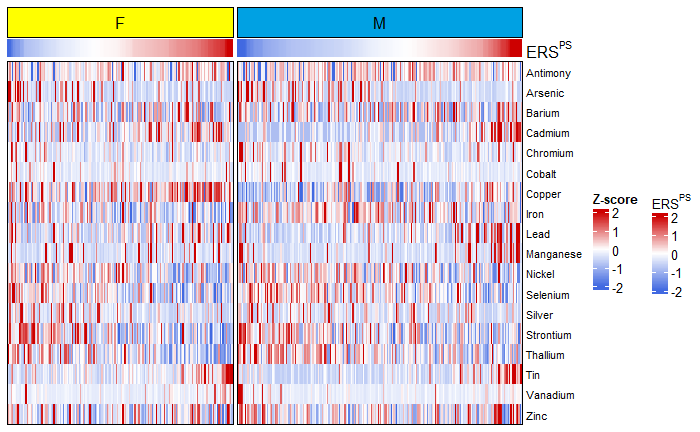

##### B. Urine

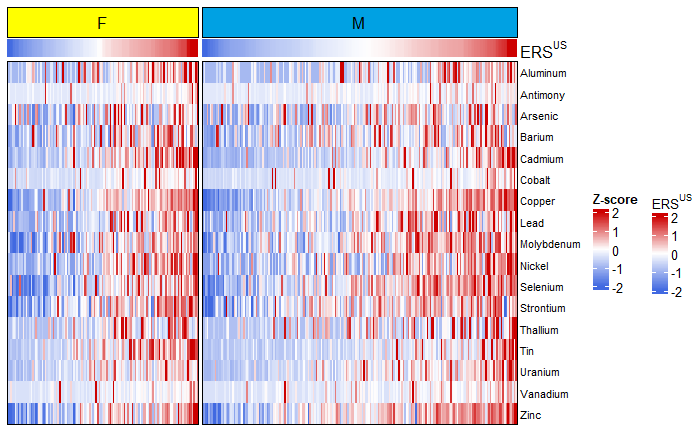

#### Supplemental Figure 10. Cohort survival analysis with amyotrophic lateral sclerosis polygenic risk score (ALS-PGS) forest plot based on plasma samples.

Cox proportional hazards survival models from plasma samples with the outcome of survival time since diagnosis (in years), the predictors are metal levels, ALS-PGS, and the interaction between metal and ALS-PGS, and adjustment covariates are age at diagnosis, sex, family history of ALS, onset segment, El-Escorial criteria, time between symptom onset and diagnosis, the first five genetic principal components, and *C9orf72* gene status (positive, negative, missing). HR 95% CI, hazard ratio and its 95% confidence interval of the metal main effect; P-value, the *p*-value of metal main effect; BH, Benjamini-Hochberg; HR 95% CI (int), hazard ratio and its 95% confidence interval of the interaction effect; P-value (int), the *p*-value of the interaction effect, red for significant interactions; %BDL, the percentage of samples below detection limit; ERS^PS^, plasma survival environmental risk score.

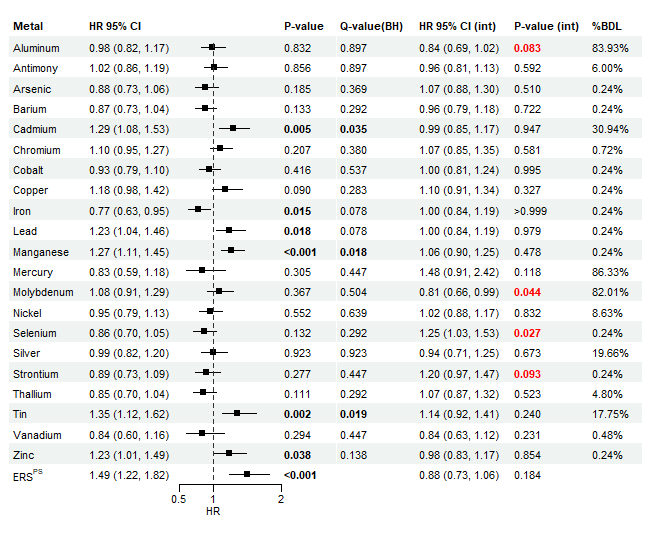

#### Supplemental Figure 11. Cohort survival analysis with metal polygenic risk score (metal-PGS) forest plot based on plasma samples.

Cox proportional hazards survival models from plasma samples with the outcome of survival time since diagnosis (in years), the predictors are metal levels, metal-PGS, and the interaction between metal and metal-PGS, and adjustment covariates are age at diagnosis, sex, family history of ALS, onset segment, El-Escorial criteria, time between symptom onset and diagnosis, the first five genetic principal components, and *C9orf72* gene status (positive, negative, missing). HR 95% CI, hazard ratio and its 95% confidence interval of the metal main effect; P-value, the *p*-value of metal main effect; BH, Benjamini-Hochberg; HR 95% CI (int), hazard ratio and its 95% confidence interval of the interaction effect; P-value (int), the *p*-value of the interaction effect, red for significant interactions; %BDL, the percentage of samples below detection limit; ERS^PS^, plasma survival environmental risk score.

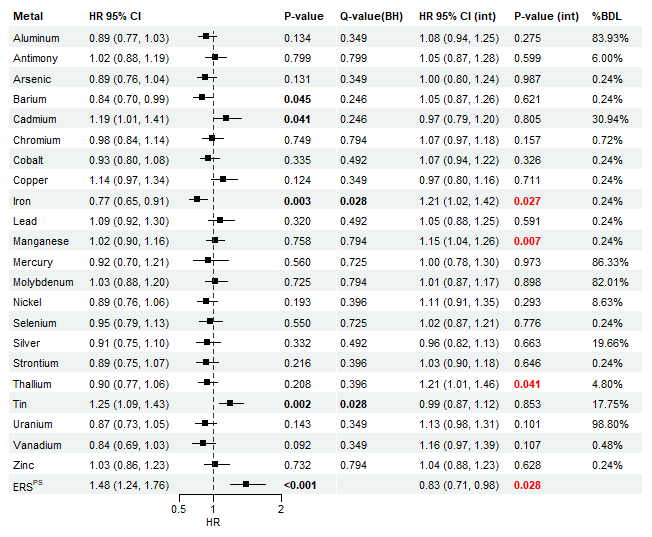

#### Supplemental Figure 12. Cohort survival analysis with amyotrophic lateral sclerosis polygenic risk score (ALS-PGS) forest plot based on urine samples.

Cox proportional hazards survival models from urine samples with the outcome of survival time since diagnosis (in years), the predictors are metal levels, ALS-PGS, and the interaction between metal and ALS-PGS, and adjustment covariates are age at diagnosis, sex, family history of ALS, onset segment, El-Escorial criteria, time between symptom onset and diagnosis, the first five genetic principal components, and *C9orf72* gene status (positive, negative, missing). HR 95% CI, hazard ratio and its 95% confidence interval of the metal main effect; P-value, the *p*-value of metal main effect; BH, Benjamini-Hochberg; HR 95% CI (int), hazard ratio and its 95% confidence interval of the interaction effect; P-value (int), the *p*-value of the interaction effect, red for significant interactions; %BDL, the percentage of samples below detection limit; ERS^US^, urine survival environmental risk score.

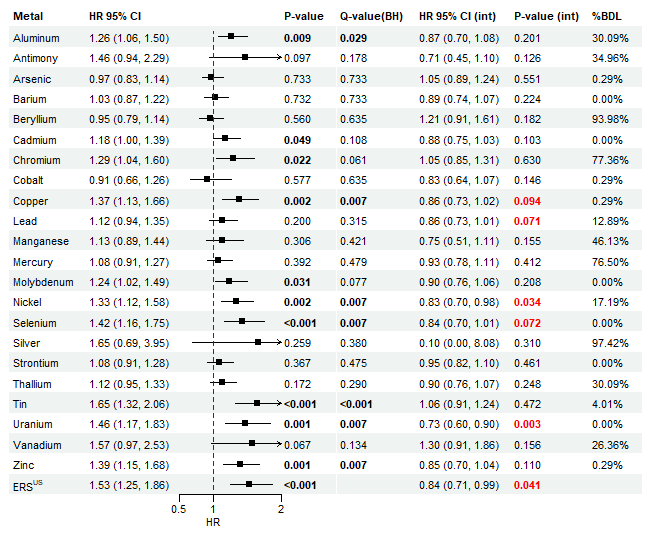

#### Supplemental Figure 13. Cohort survival analysis with metal polygenic risk score (metal-PGS) forest plot based on urine samples.

Cox proportional hazards survival models from urine samples with the outcome of survival time since diagnosis (in years), the predictors are metal levels, metal-PGS, and the interaction between metal and metal-PGS, and adjustment covariates are age at diagnosis, sex, family history of ALS, onset segment, El-Escorial criteria, time between symptom onset and diagnosis, the first five genetic principal components, and *C9orf72* gene status (positive, negative, missing). HR 95% CI, hazard ratio and its 95% confidence interval of the metal main effect; P-value, the *p*-value of metal main effect; BH, Benjamini-Hochberg; HR 95% CI (int), hazard ratio and its 95% confidence interval of the interaction effect; P-value (int), the *p*-value of the interaction effect, red for significant interactions; %BDL, the percentage of samples below detection limit; ERS^US^, urine survival environmental risk score.

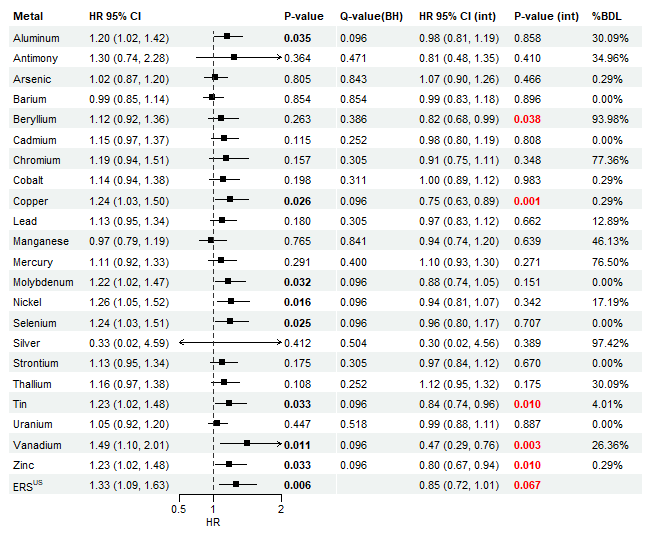

#### Supplemental Figure 14. Plasma and occupational exposure network analysis.

Associations between self-reported occupational metal exposures from participants’ most recent job and plasma metals estimated by the partial least squares method. Red, positive association; Blue, negative association.

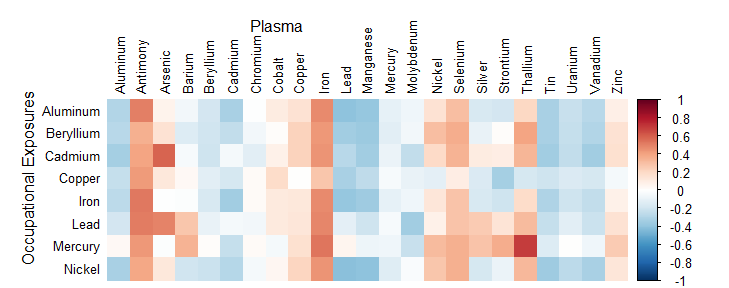

#### Supplemental Figure 15. Plasma and non-occupational exposure network analysis.

Associations between self-reported non-occupational metal exposures and plasma metals estimated by the partial least squares method. Red, positive association; Blue, negative association.

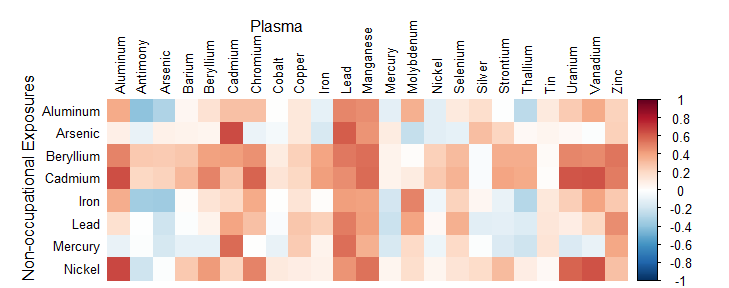

#### Supplemental Figure 16. Urine and occupational exposure network analysis.

Associations between self-reported occupational metal exposures from participants’ most recent job and urine metals estimated by partial least squares method. Red, positive association; Blue, negative association.

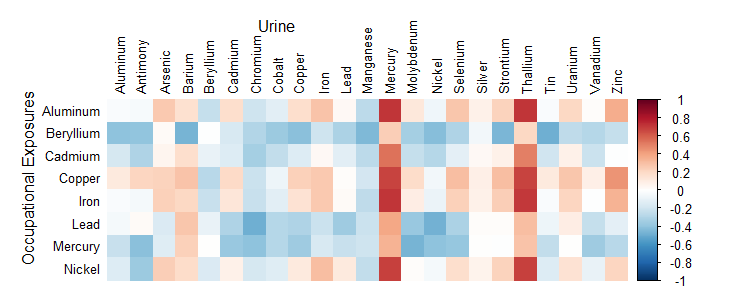

#### Supplemental Figure 17. Urine and non-occupational exposure network analysis.

Associations between self-reported non-occupational metal exposures and urine metals estimated by partial least squares method. Red, positive correlation; Blue, negative correlation.

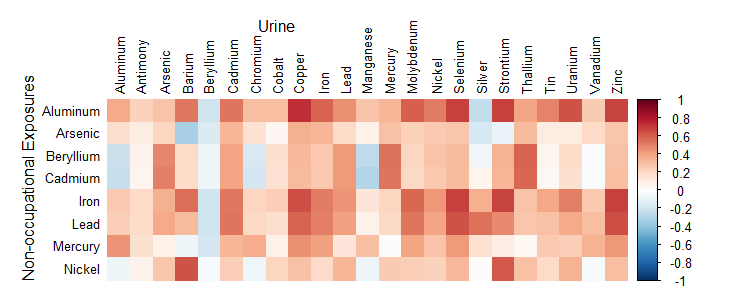
